# Low incidence of adverse events or construct failure of a novel high-strength No.2 round suture in rotator cuff repair: An IDEAL Stage 2a assessment retrospective cohort analysis

**DOI:** 10.1101/2024.08.19.24312206

**Authors:** Cooper Moody, Corey Scholes, Manaal Fatima, Kevin Eng, Graeme Brown, Richard S Page

**Affiliations:** Barwon Health, Geelong, Victoria, Australia; EBM Analytics, Sydney, New South Wales, Australia; Geelong Orthopaedics, Geelong, Victoria, Australia; Barwon Centre for Orthopaedic Research and Education (B-CORE), Deakin University, Victoria, Australia; St John of God Hospital, Geelong, Victoria, Australia

**Author notes:** **Corresponding author:** Kevin Eng, Geelong Orthopaedics, Level 2/83 Myers St, Geelong VIC 3220, Australia.

**Keywords:** Rotator cuff tear, rotator cuff repair, suture, Dynacord, patient reported outcome measure, WORC, QuickDASH, IDEAL framework

## Abstract

**Background:** Despite technical and material improvements in rotator cuff repair (RCR) clinical and radiological failure remains common. Following suture fixation, tension and footprint compression decrease from time zero. A novel suture (Dynacord, Depuy Synthes) has been designed to shorten when submerged in liquid to maintain tension and increase repair construct security.

**Methods:** A retrospective cohort analysis was performed on the PRULO (Patient Reported Outcomes in Upper Limb Surgery) registry for 12 months follow up after RCR using this suture regarding all cause failure, rates of common complications, Quick Disability of the Arm, Shoulder and Hand (QuickDASH), and Western Ontario Rotator Cuff Index (WORC). Summary statistics were generated for patient characteristics and patient-reported outcome measures (PROMs) analysed using multiple imputation and a linear model to assess changes over 12 months follow up.

**Results:** A cohort of 236 cases was included for analysis. Complication rates and functional improvements were comparable to literature on similar sutures. At 12 months follow up, all-cause failure occurred in 12% of cases, and mean scores for QuickDASH decreased by 37 and WORC increased by 44, both of which surpass the minimum clinically important difference. Our observed rates of complications are as follows: Infection 2.1%, stiffness/capsulitis 11% and retear 12%.

**Conclusion:** The novel suture demonstrated favourable safety and efficacy profiles, with outcomes comparable to those published for commonly used sutures. This study through an IDEAL 2a framework for surgical innovation highlights this suture as safe, effective in mitigating common failure mechanisms and having satisfactory outcomes in RCR.

## Introduction

Rotator cuff tears (RCT) account for up to 70% of shoulder related clinician visits (Rees, 2008) and are present in at least 60% of people in the United States over 60 years of age (Kukkonen et al., 2015). Surgical management of RCT is increasing in frequency (Colvin et al., 2012; Yanik et al., 2021). Rotator cuff repair (RCR) has been shown to provide benefits in Quality-Adjusted Life Years and net societal cost saving (Mather et al., 2013). Patients with a radiologically intact surgical rotator cuff repair at one year post procedure have shown significantly improved pain, function and disability (Lambers Heerspink et al., 2015).

RCR failure has been observed at rates between 7.5% and 94% (Chillemi et al., 2018; Chona et al., 2017; Jost et al., 2000)(Ren et al., 2019)(Chillemi et al., 2018; Chona et al., 2017; Jost et al., 2000). Failure of RCR can broadly be defined as radiological or direct intra-operative visualisation of rotator cuff retear or construct failure, significant complications, worsening of pre-operative function or a lack of reasonably perceived functional improvement (Desmoineaux, 2019). Construct failure mechanisms include suture anchor pullout from bone, pull-through of suture through tendon (‘cheese wiring’), knot failure, suture laxity or creep, suture rupture and failure of tendon to bone healing (Borbas et al., 2021; Hurwit et al., 2014; Lapner et al., 2022; Savage et al., 2023). An ideal suture design to mitigate these would exhibit low abrasiveness to tendon, a high load to rupture, maintenance of high tension once tied, robust knot security ((Borbas et al., 2021; Hurwit et al., 2014; Lapner et al., 2022)), be of a low profile and produce minimal debris formation at contact points with the anchor (Lovric et al., 2018; Savage et al., 2023).

The high strength No. 2 suture (Dynacord, Depuy Synthes) has a novel design with an outer sheath of ultra-high molecular weight polyethylene (UHMWPE), an inner polyester sheath and a silicone and sodium chloride filled core. When submerged in fluid the sodium chloride dissolves and leaves a tri-porous structure filled with water. This causes the suture braid to expand radially and shorten axially, which maintains the suture tension (Pastor et al., 2024). In laboratory-based testing this suture demonstrated increased force to knot failure (Ensminger et al., 2021), reduced tendon to bone gap formation, decreased throws required for knot security and maintained greater compression force over the footprint area when compared to a static suture.

Static suture tensile strength has been well investigated. What has not been thoroughly explored is how creep and subsequent tension loss post suture time zero fixation can be negated. This novel biomaterials approach to RCR is in the idea and development stage of the IDEAL framework for surgical innovation (Mculloch et. al 2013). Crucial to this early investigation is analysis of proof of concept, safety and efficacy in a heterogeneous cohort. This analysis may act as a framework for a future study with a larger cohort, greater follow up and further functional and clinical outcome measures.

The aim of this study was to describe the proof of concept, safety and clinical outcomes in patients receiving rotator cuff repair with a novel high-strength No. 2 round suture with a minimum of 12 months follow up.

## Methods

### Study Design

Retrospective cohort analysis retrieved from a single-centre, prospective clinical registry, with institutional ethics approval. The RECORD guidelines (Benchimol et al., 2015) were observed in the reporting of this study (Supplementary 1).

### Registration and Ethics

The Patient Registry of Upper Limb pathology Outcomes (PRULO) registry is approved for research output by a nationally-recognised Human Research Ethics Committee (Barwon Health, Project ID 49184, Reference 19/70) and registered on the Australian New Zealand Clinical Trials Registry (ACTRN12619000770167).

### Setting

PRULO is a multi-cohort, prospective observational, clinical quality registry collating clinical data and patient-reported outcomes (PROMs) for patients presenting to a specialist orthopaedic clinic (Scholes et al., 2023a). PRULO captures patient reported outcomes including the abbreviated Disabilities of the Arm, Shoulder and Hand (QuickDASH), Western Ontario Rotator Cuff (WORC) Index as well as clinical and radiological data. Data points are recorded at initial practice registration, after initial consultation, intraoperatively, as well as three, six, 12 and 24 months following definitive intervention. Patient subgroups (cohorts) include conditions affecting predominantly the rotator cuff (tear, tendinopathy), conditions associated with glenohumeral instability, as well as all other conditions presenting in the shoulder, according to the surgeon-generated diagnosis.

### Participants and Grouping

The cohort of patients undergoing shoulder surgery involving fixation with a No. 2 round suture (Dynacord, Depuy-Mitek, USA) were extracted by matching product stock keeping units (SKU), retrieved from packaging information retained in surgery. The list of included SKUs are detailed in Supplementary 2. Any patient undergoing repair with the novel suture was included, independent of patient or tear characteristics.

### Surgical Technique

Three consultant orthopaedic surgeons were involved. All surgeons utilised double row transosseous equivalent repairs. Two surgeons utilised an all arthroscopic technique in beach chair position and one surgeon utilised a mini open approach in lateral position. The novel suture was utilised with a combination of PEEK (Polyether-Ether-Ketone) and BR (Biocryl-Rapide) suture anchors depending on surgeon preference. Free suture was utilised when margin convergence was performed. Adjunct procedures were sometimes performed including biceps tenodesis or tenotomy, subacromial decompression and clavicle resection amongst others. Postoperatively patients had their shoulder immobilised in a sling for six weeks and restricted to passive shoulder motion. From 6-12 weeks, active assisted range of motion was allowed and strengthening exercises added at 12 weeks.

### Outcomes

The variables included in the analysis were retrieved from the registry core dataset as detailed previously (Scholes et al., 2023a). These included patient demographics characteristics, treatment details, adverse events, procedure survival and patient-reported outcomes as described in Supplementary 2.

### Statistical Analysis

Descriptives for continuous variables of patient characteristics were reported using medians and interquartile ranges (IQR), counts and proportions were reported for categorical variables (Supplementary 2). Complications were retrieved for each treatment record where present and descriptive statistics generated for all complications observed. For time to event analysis for intraoperative and postoperative adverse events, duration (*surgery date* to *end date*) was summarised with mean and standard deviation and visualised with ridge plots. A Kaplan-Meier survival curve (with 95% confidence intervals) with procedure *failure* as the event of interest was plotted. A linear model was applied to each imputed dataset forQuickDASH and WORC Index, with *Timepoint* as the primary predictor and *Age at surgery* and *Sex* included as covariates. The results from each model were pooled for presentation including coefficients, 95% confidence intervals and p-values. P-values for Sex and Timepoint were reported for comparisons against a reference level. Pooled predictions for the PROMs were calculated from linear models across the multiple-imputed datasets (Arel-Bundock, 2023) and plotted across *Timepoint*.

## Results

Of the 945 surgical cases eligible for analysis (Supplementary 2), 236 cases involved the suture of interest. Median follow up was 24 months (IQR 15 – 33 months) with a minimum of 12 months.

### Patient characteristics

The studied cohort comprised 25% females, had a mean age of 58, and 36% of patients had surgery on their non-dominant side. The mean time from the initial consult to surgery was 20 weeks (Table 1). Of the sample, 92% of patients had full thickness tears of predominantly medium size (Table 2). Crescentic shaped tears were most common. Surgery was performed predominantly arthroscopically in beach chair position using anatomic double-row repair of the supraspinatus in isolation using knotted fixation in the majority of cases (Supplementary 2).

**Table 1:**
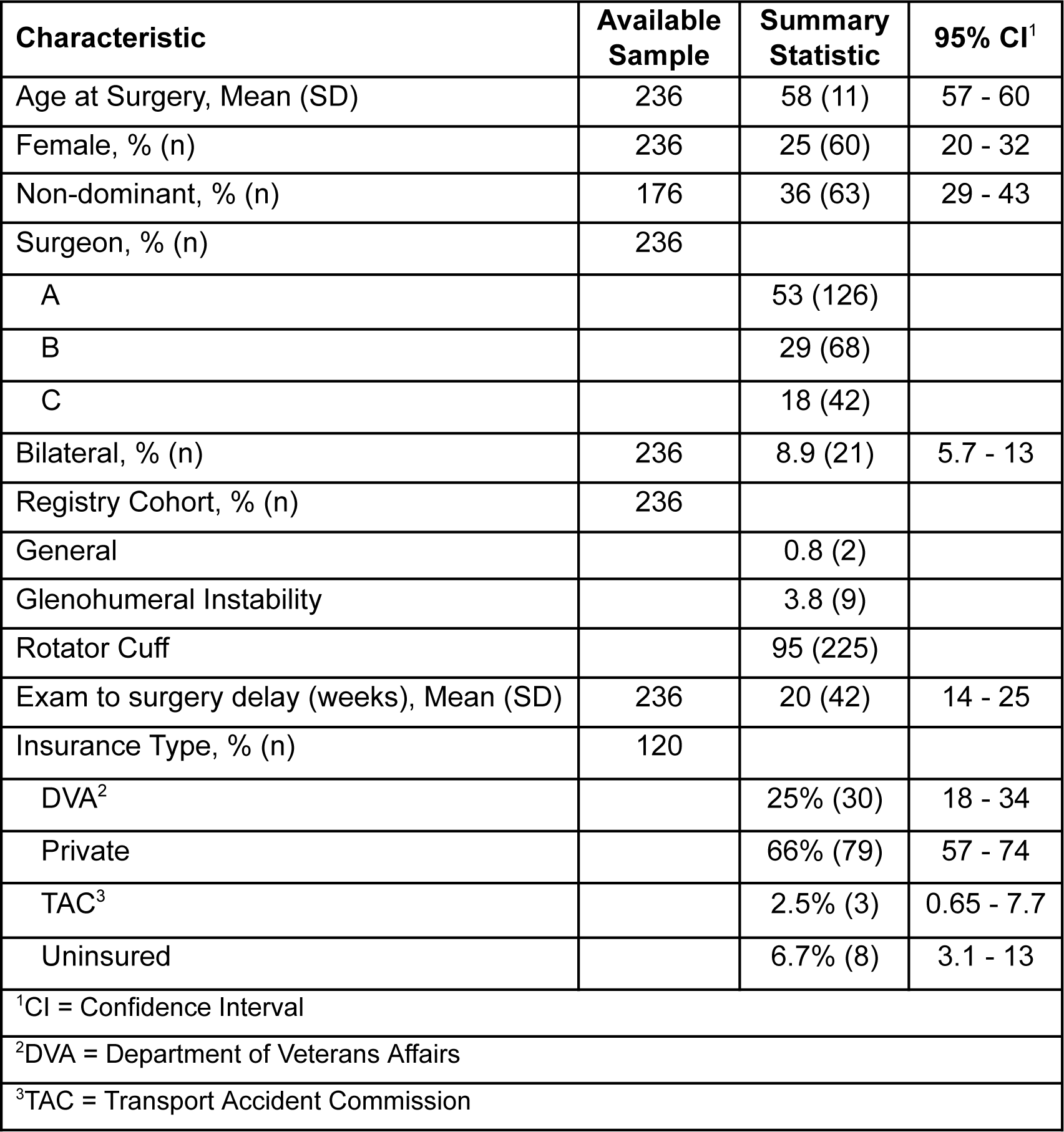
Summary of patient characteristics for the cohort.

| **Characteristic** | **Available Sample** | **Summary Statistic** | **95% CI<sup>1</sup>** |
| --- | --- | --- | --- |
| Age at Surgery, Mean (SD) | 236 | 58 (11) | 57 - 60 |
| Female, % (n) | 236 | 25 (60) | 20 - 32 |
| Non-dominant, % (n) | 176 | 36 (63) | 29 - 43 |
| Surgeon, % (n) | 236 |  |  |
| A |  | 53 (126) |  |
| B |  | 29 (68) |  |
| C |  | 18 (42) |  |
| Bilateral, % (n) | 236 | 8.9 (21) | 5.7 - 13 |
| Registry Cohort, % (n) | 236 |  |  |
| General |  | 0.8 (2) |  |
| Glenohumeral Instability |  | 3.8 (9) |  |
| Rotator Cuff |  | 95 (225) |  |
| Exam to surgery delay (weeks), Mean (SD) | 236 | 20 (42) | 14 - 25 |
| Insurance Type, % (n) | 120 |  |  |
| DVA<sup>2</sup> |  | 25% (30) | 18 - 34 |
| Private |  | 66% (79) | 57 - 74 |
| TAC<sup>3</sup> |  | 2.5% (3) | 0.65 - 7.7 |
| Uninsured |  | 6.7% (8) | 3.1 - 13 |
| <sup>1</sup>CI = Confidence Interval |  |  |  |
| <sup>2</sup>DVA = Department of Veterans Affairs |  |  |  |
| <sup>3</sup>TAC = Transport Accident Commission |  |  |  |

**Table 2:**
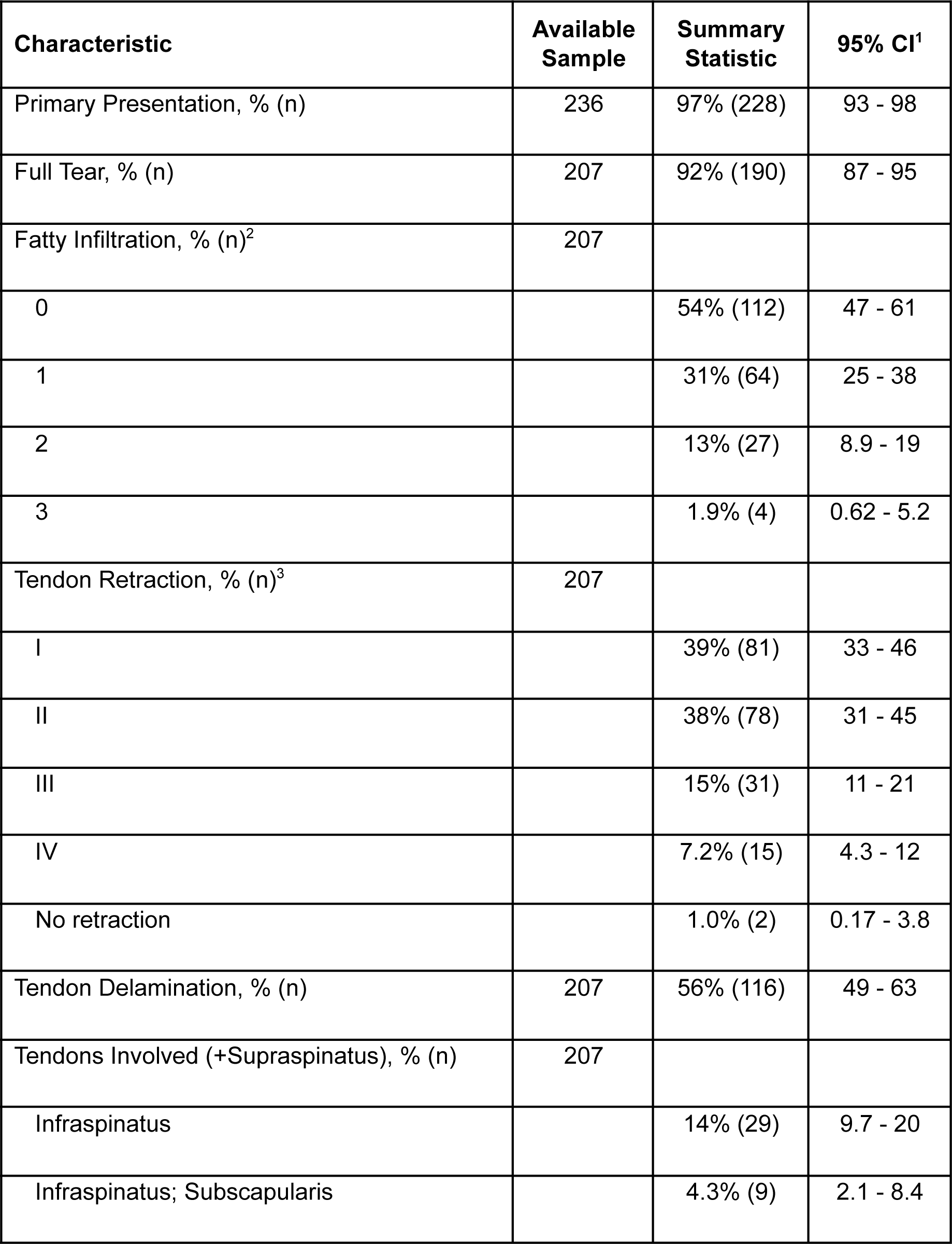
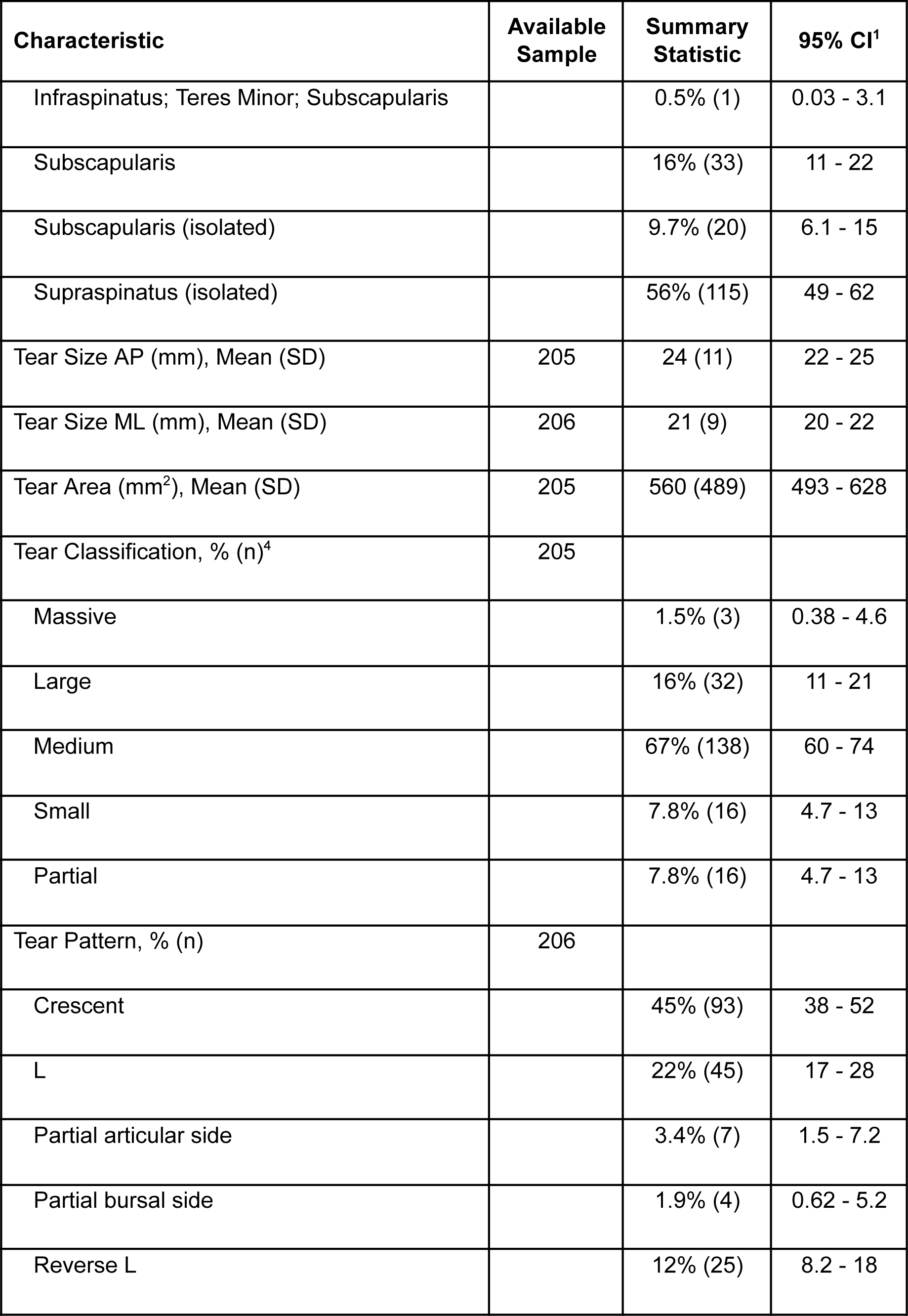
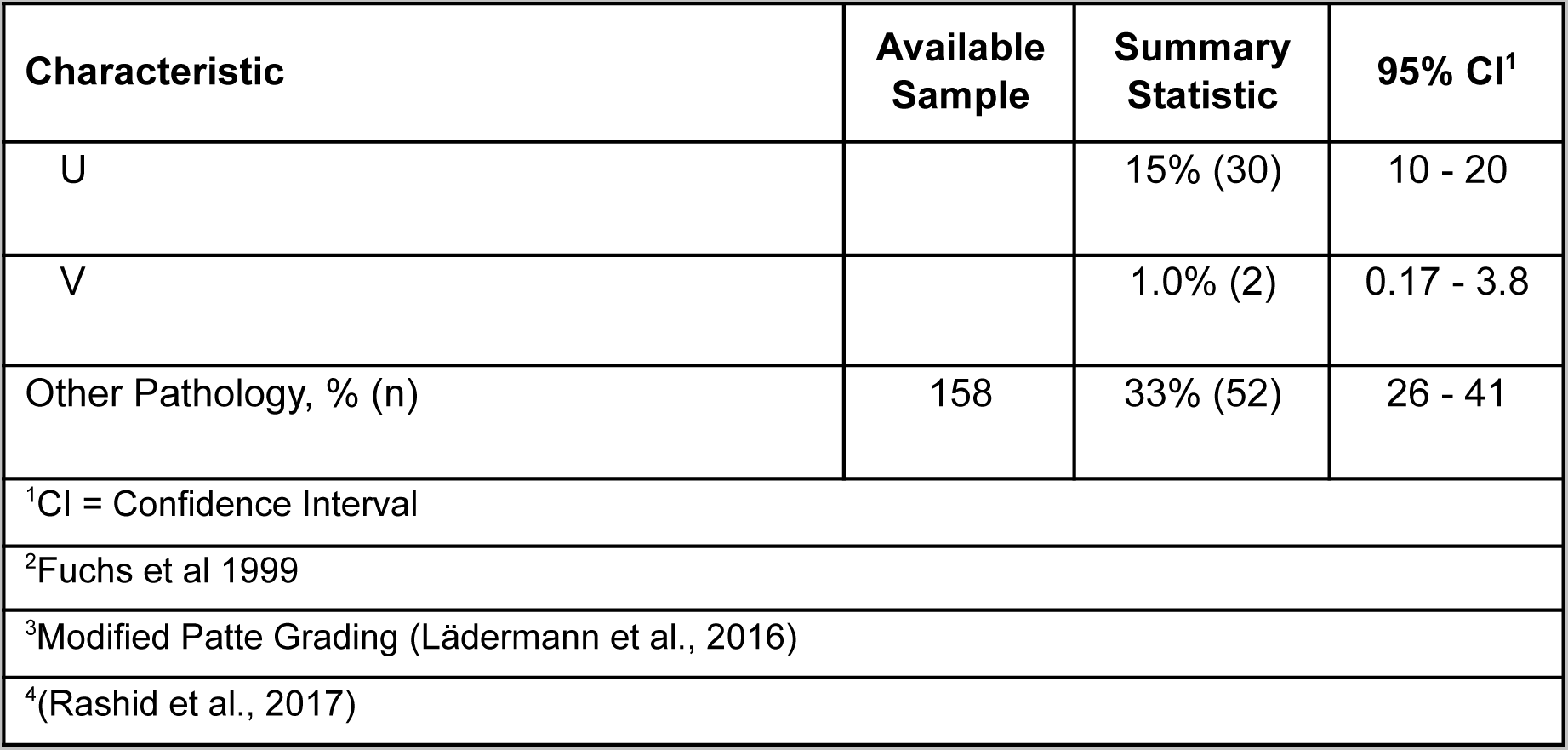
Summary of tear characteristics of rotator cuff repair cases.

### Complications

Complications were observed in 29% (N=77) of the cohort. The most common complications post operatively were stiffness and failure of repair (Table 3). Twenty eight patients had a retear (N = 28 (12)%).Twenty six had capsulitis. (N = 26 (11)%, 95%CI 7.5 – 16%). Two cases underwent implant removal due to loosening or infection (N = 2, (0.8)%, 95%CI 0.15 – 3.4%).

**Table 3:**
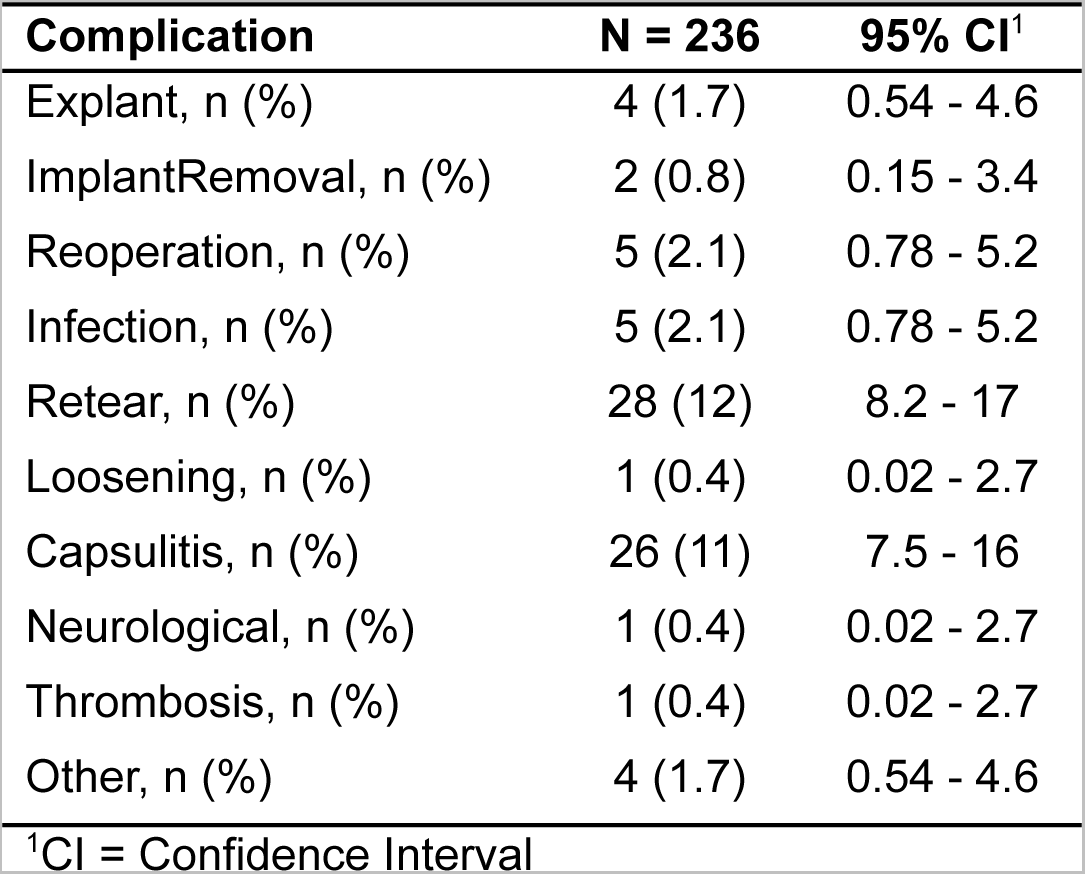
Incidence of complications (and reintervention) identified in the study sample from minimum of 30 days after surgery to latest follow up.

The majority of adverse events were of low severity (Grade I), without requiring additional intervention, or Grade III (Table 4). There were four cases classed as Grade IV – one retear converted to total shoulder replacement, three cases of retear deemed irreparable and one chronic hemi-diaphragm palsy.

**Table 4:**
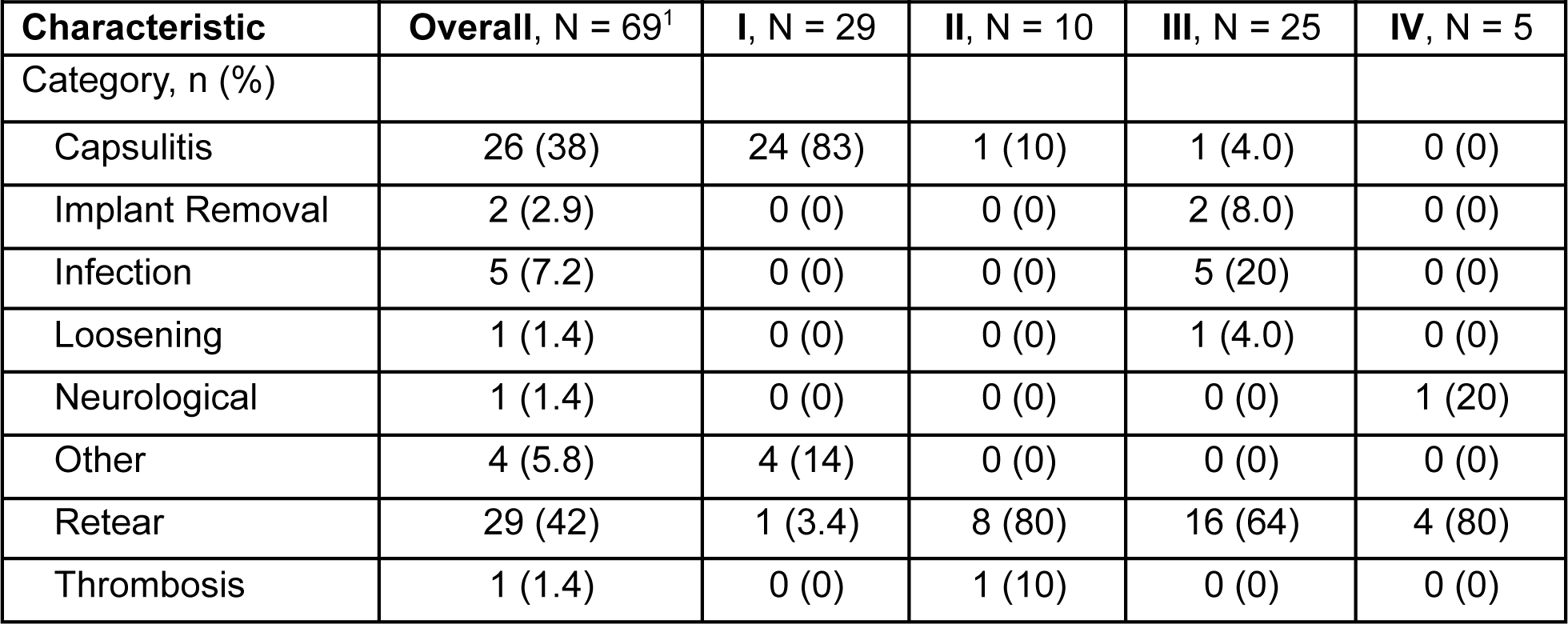
Cross tabulation of adverse event category with modified Sink grade for severity.

### Time to Event

Time to event analysis revealed retear-free survival of 87.8% (95%CI 82.9 – 91.4) at 12 months. In addition, different incidence trajectories were observed for adverse events, when taking into account retear and implant removal as competing risks (Figure 1). In addition, infection peaked at 0.4% (95%CI 0.1 – 3.0) at 4 weeks follow up.

**Figure 1:**
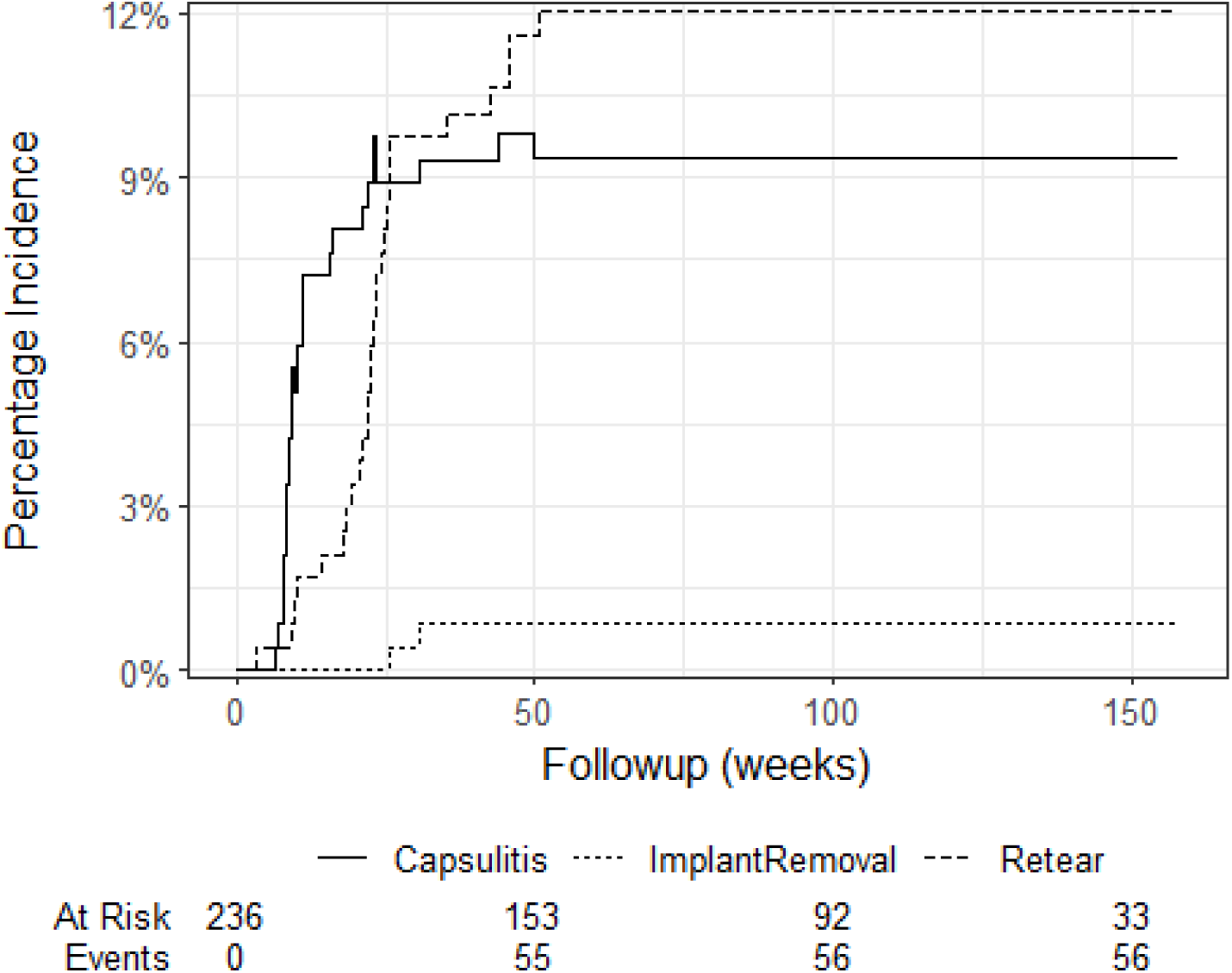
Cumulative incidence of capsulitis in the presence of competing risks (implant removal and tendon retear)

### Patient-reported outcomes

Model-predicted scores displayed a reduction in QDASH total score median of 30 points at 6 months and 37 points at 12 months (Table 5a). Similarly the median normalised WORC Index improved by 35 and 44 points at 6 and 12 months respectively (Table 5b). Distribution plots of the model-predicted scores demonstrated considerable between-patient variation in response trajectories (Figure 2).

**Figure 2:**
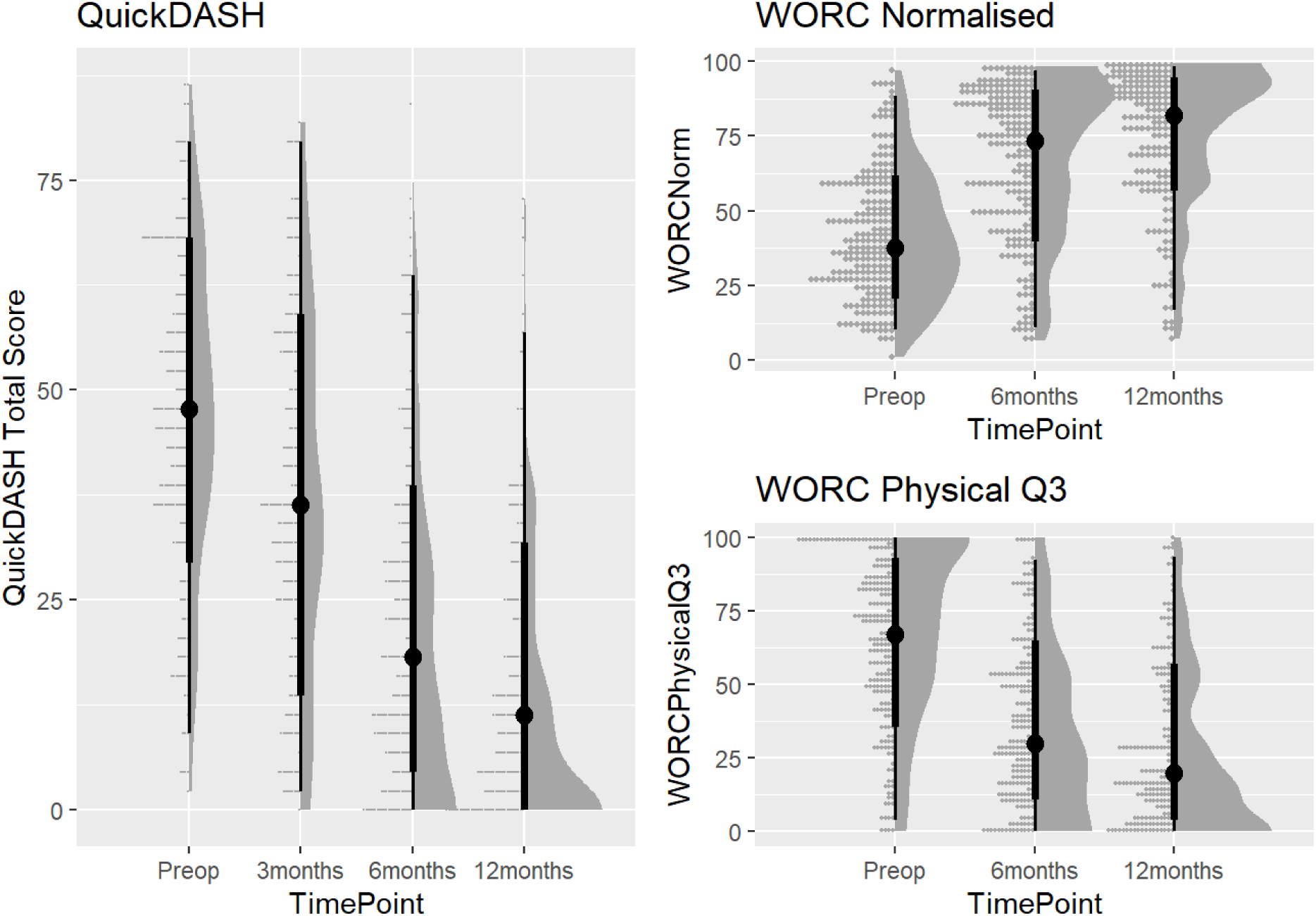
Trajectories of model-predicted QuickDASH (left), WORC Index Normalised (top right) and WORC Physical Q3 (bottom right) across time points.

**Table 5a:**
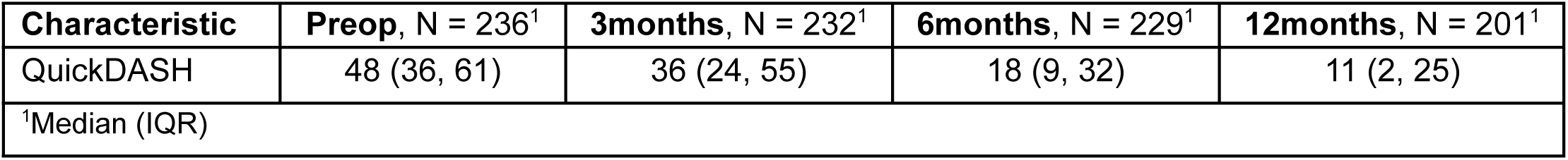
Summary of model-predicted QuickDASH by TimePoint.

| Characteristic | Preop, N = 236<sup>1</sup> | 3months, N = 232<sup>1</sup> | 6months, N = 229<sup>1</sup> | 12months, N = 201<sup>1</sup> |
| --- | --- | --- | --- | --- |
| QuickDASH | 48 (36, 61) | 36 (24, 55) | 18 (9, 32) | 11 (2, 25) |
| <sup>1</sup>Median (IQR) |  |  |  |  |

**Table 5b:**
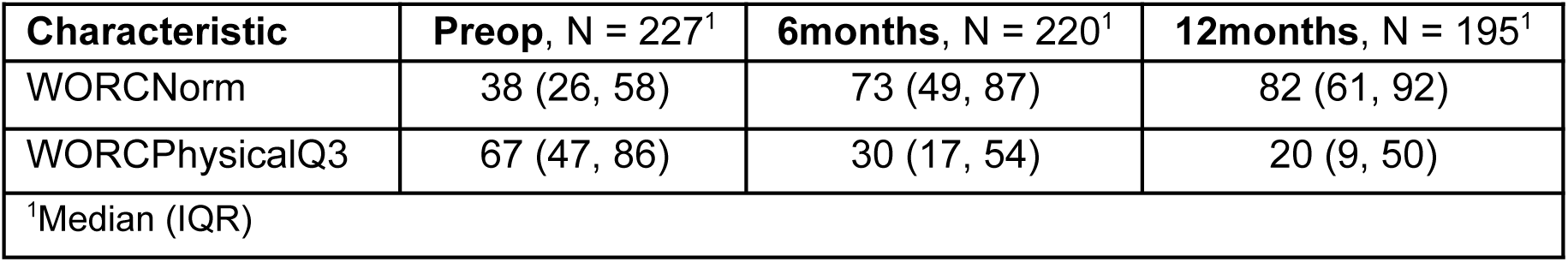
Summary of model-predicted WORC by TimePoint.

## Discussion

This study aimed to describe the proof of concept, safety and clinical outcomes at a minimum of 12 months followup in patients who received RCR with a novel high-strength No. 2 round suture. The key findings were a low incidence of postoperative complications and improvement in PROMs aligned with the expected trajectory for recovery. To our knowledge there is no existing clinical literature on outcomes using this specific suture for RCR in vivo. Median follow-up for the present cohort extended to 24 months, but PROMs data was available for a minimum of 12 months.

The patient population studied here is similar to previously published studies (Maher et al., 2022), with some differences. The majority of patients presented with a medium size tear (67%), while the remaining third of the cohort ranged between small to massive tears. Patients with larger tears have a higher rate of retear in general (Zhao et al., 2021). Tear pattern was most commonly crescentic, but six other patterns accounted for 55% of the sample. The cohort was largely male and privately funded, two factors which may lead to better clinical and patient-reported outcomes (Mandalia et al., 2023). The suture was also used in 29 combinations of anchors (Supplementary 2). However, this variability may aid in the generalisability of the present analysis.

Complications and adverse events occurred in 29% of the cohort, with an all-cause failure (retear) rate of 12% at 12 months. For the same follow-up period, previous meta-analyses have reported failure rates of 12-14% (with early versus delayed rehabilitation) (Mazuquin et al., 2021), 20% (Boksh et al., 2022) and up to 43% (Rashid et al., 2017). One randomised control trial reported radiographically evident retears in 31-34% of their patients (undergoing early versus delayed postoperative motion protocols) 6 months after RCR (Mazzocca et al., 2017). Construct failure or requirements for revision RCR are documented within the first 6 months after surgery in most studies (Longo et al., 2021).

The other common adverse event reported in this cohort was stiffness, observed in 11% of patients. Shoulder stiffness is a common and considerable complication after arthroscopic RCR, with a prevalence ranging between 3-23% (Saade et al., 2023), and calculated at 6.4% in one systematic review, regardless of tear type (Baumann et al., 2023). Reported *deep* infection rates following open or mini-open rotator cuff repair range from 0.3-1.9% (Athwal et al., 2007) with previous work from our group observing a rate of 1.2% after mini-open repair (Asaid et al., 2018), whereas the rate for arthroscopic repair ranges from 0.3-0.9% (Stone et al., 2023)The infection rate from the study was marginally higher at 2.1%, observed in five cases, which is likely an artefact of the sample size (illustrated by wide confidence intervals 0.78 – 5.2%) and between-study variations in definitions. The rates of complication observed in this study are comparable to those in published evidence, and importantly no novel complications, specific to the suture of interest, were observed.

Candidates for RCR present with a typical baseline QuickDASH score of 51.9 ± 20.2, and a baseline WORC of 32.0 ± 18.3 (Macdermid et al., 2015). The mean baseline scores for this cohort were comparable – QuickDASH (48) and WORC (38) respectively. QuickDASH scores at 6 months in this cohort were superior to other studies (Abufoul et al., 2023; Macdermid et al., 2015), and at 12 months, they were comparable to some reports (Leow et al., 2022; Zwolak et al., 2022) and better than others (Correia et al., 2022). In one systematic review, DASH scores improved in patients undergoing rotator cuff repair from 50.2 preoperatively, to 19 at a minimum of 12 months postoperatively (Unger et al., 2019). When comparing DASH scores to QuickDASH, similar score estimates can be expected, though QuickDASH is typically 1-3 points higher in an RCR cohort (Macdermid et al., 2015).

The trajectory of WORC scores in this cohort were most similar to a systematic review (Sahoo et al., 2021), with baseline scores of 38 and 12 month scores of 82. At 6 months, WORC scores in this study were superior to some (Macdermid et al., 2015) and inferior to others (Sheps et al., 2019). The minimum clinically important difference (MCID) for a moderate improvement in QuickDASH is 15.9 points (Franchignoni et al., 2014), and 13.1 for the WORCNorm (Jones et al., 2020), which has been surpassed by the lower confidence bound in the imputed model for both 6 months and 12 months of follow up.

As this study is based on a clinical registry, there are inherent risks for biases, including selection bias, information bias, classification bias and process drift. Reporting and follow up may be hindered by competency, language or technological barriers, and the present cohort may not be representative of a general sample, as evidenced by inclusion of majority males, above-baseline scores and privately funded patient status (Scholes et al., 2023a).

There is a lack of a comparator group in this retrospective cohort analysis. The group is heterogeneous in pathology, for instance tear size and shape. Comparison between studies with respect to PROMs is also complicated by a lack of standardisation of outcomes reporting in patients undergoing RCR (Ardebol et al., 2023). Some confounding factors may also be introduced in that this sample is exclusively from three surgeons in a regional private practice in Australia.

Declarations regarding success or failure at this stage (IDEAL framework Stage 2a) may not represent the full spectrum of outcomes that could be seen in a larger study with longer follow-up, but such studies may not be plausible without the promising results of short-term observational studies such as those presented here. Future studies may address radiological analysis of the suture and anchors utilised, as well as clinical results over the longer term.

## Conclusion

These findings highlight the potential of the Dynacord No. 2 suture as a safe and viable solution in RCR. The clinical improvements observed in WORC and QuickDASH scores surpassed the MCID and displayed non-inferiority to existing studies. Complications including all-cause failure were comparable to previous studies in both nature and incidence. Importantly, there were no novel complications. The study presents an idea and development stage of the IDEAL framework for surgical innovation, and provides evidence to warrant larger-scale studies or randomised control trials to further evaluate the efficacy of the suture in a more diverse population.

## Data Availability

All data produced in the present study are available upon reasonable request to the authors

## Acknowledgements

The authors would like to acknowledge the participation of the patients in the PRULO registry and contributions of the staff at Geelong Orthopaedics for the conduct of the study. The authors would also like to thank Milad Ebrahimi (EBM Analytics) and other team members for assistance with data collection and extraction for this study.

## References

1. Abufoul, R., Gavish, L., & Haddad, M. (2023). Photobiomodulation self-treatment at home after rotator cuff arthroscopic repair accelerates improvement in pain, functionality, and quality of life: A double-blind, sham-controlled, randomized clinical trial. Lasers in Surgery and Medicine, 55(7), 662–673.

2. Ardebol, J., Ghayyad, K., Hwang, S., Pak, T., Menendez, M. E., & Denard, P. J. (2023). Patient-reported outcome tools and baseline scores vary by country and region for arthroscopic repair of massive rotator cuff tears: a systematic review. *JSES Reviews*, Reports, and Techniques, 3(3), 312–317.

3. Arel-Bundock, V. (2023). _marginaleffects: Predictions, Comparisons, Slopes, Marginal Means, and Hypothesis Tests_(Version 0.15.0) [R package]. https://marginaleffects.com/

4. Asaid, R., Eng, K., Brown, G., & Page, R. (2018). Long-term outcomes after infected mini-open rotator cuff repair: results of a 10-year review. Journal of Shoulder and Elbow Surgery / American Shoulder and Elbow Surgeons … [et Al.], 27(4), 751–755.

5. Athwal, G. S., Sperling, J. W., Rispoli, D. M., & Cofield, R. H. (2007). Deep infection after rotator cuff repair. Et Al [Journal of Shoulder and Elbow Surgery], 16(3), 306–311.

6. Baumann, A. N., Oleson, C., Curtis, D. P., Indermuhle, T., & Leland, J. M., 3rd. (2023). The Incidence of Postoperative Shoulder Stiffness After Arthroscopic Rotator Cuff Repair: A Systematic Review. Cureus, 15(4), e37199.

7. Boksh, K., Haque, A., Sharma, A., Divall, P., & Singh, H. (2022). Use of Suture Tapes Versus Conventional Sutures for Arthroscopic Rotator Cuff Repairs: A Systematic Review and Meta-analysis. The American Journal of Sports Medicine, 50(1), 264–272.

8. Borbas, P., Fischer, L., Ernstbrunner, L., Hoch, A., Bachmann, E., Bouaicha, S., & Wieser, K. (2021). High-Strength Suture Tapes Are Biomechanically Stronger Than High-Strength Sutures Used in Rotator Cuff Repair. Sports Medicine, Arthroscopy, Rehabilitation, Therapy & Technology: SMARTT, 3(3), e873–e880.

9. Chillemi, C., Dei Giudici, L., Mantovani, M., Osimani, M., & Gumina, S. (2018). Rotator cuff failure after surgery: an all-arthroscopic transosseous approach. Musculoskeletal Surgery, 102(Suppl 1), 3–12.

10. Chona, D. V., Lakomkin, N., Lott, A., Workman, A. D., Henry, A. C., Kuntz, A. F., Huffman, G. R., & Glaser, D. L. (2017). The timing of retears after arthroscopic rotator cuff repair. Journal of Shoulder and Elbow Surgery / American Shoulder and Elbow Surgeons … [et Al.], 26(11), 2054–2059.

11. Colvin, A. C., Egorova, N., Harrison, A. K., Moskowitz, A., & Flatow, E. L. (2012). National trends in rotator cuff repair. The Journal of Bone and Joint Surgery. American Volume, 94(3), 227–233.

12. Correia, F. D., Molinos, M., Luís, S., Carvalho, D., Carvalho, C., Costa, P., Seabra, R., Francisco, G., Bento, V., & Lains, J. (2022). Digitally Assisted Versus Conventional Home-Based Rehabilitation After Arthroscopic Rotator Cuff Repair: A Randomized Controlled Trial. American Journal of Physical Medicine & Rehabilitation / Association of Academic Physiatrists, 101(3), 237–249.

13. Desmoineaux, P. (2019). Failed rotator cuff repair. Orthopaedics & Traumatology, Surgery & Research: OTSR, 105(1S), S63–S73.

14. Ensminger, W. P., McIff, T., Vopat, B., Mullen, S., & Schroeppel, J. P. (2021). Mechanical Comparison of High-Strength Tape Suture Versus High-Strength Round Suture. Sports Medicine, Arthroscopy, Rehabilitation, Therapy & Technology: SMARTT, 3(5), e1525–e1534.

15. Franchignoni, F., Vercelli, S., Giordano, A., Sartorio, F., Bravini, E., & Ferriero, G. (2014). Minimal clinically important difference of the disabilities of the arm, shoulder and hand outcome measure (DASH) and its shortened version (QuickDASH). The Journal of Orthopaedic and Sports Physical Therapy, 44(1), 30–39.

16. Hurwit, D., Fanton, G., Tella, M., Behn, A., & Hunt, K. J. (2014). Viscoelastic properties of common suture material used for rotator cuff repair and arthroscopic procedures. Arthroscopy: The Journal of Arthroscopic & Related Surgery: Official Publication of the Arthroscopy Association of North America and the International Arthroscopy Association, 30(11), 1406–1412.

17. Jones, I. A., Togashi, R., Heckmann, N., & Vangsness, C. T., Jr. (2020). Minimal clinically important difference (MCID) for patient-reported shoulder outcomes. Journal of Shoulder and Elbow Surgery / American Shoulder and Elbow Surgeons … [et Al.], 29(7), 1484–1492.

18. Jost, B., Pfirrmann, C. W., Gerber, C., & Switzerland, Z. (2000). Clinical outcome after structural failure of rotator cuff repairs. The Journal of Bone and Joint Surgery. American Volume, 82(3), 304–314.

19. Kukkonen, J., Joukainen, A., Lehtinen, J., Mattila, K. T., Tuominen, E. K. J., Kauko, T., & Äärimaa, V. (2015). Treatment of Nontraumatic Rotator Cuff Tears: A Randomized Controlled Trial with Two Years of Clinical and Imaging Follow-up. The Journal of Bone and Joint Surgery. American Volume, 97(21), 1729–1737.

20. Lambers Heerspink, F. O., van Raay, J. J. A. M., Koorevaar, R. C. T., van Eerden, P. J. M., Westerbeek, R. E., van’t Riet, E., van den Akker-Scheek, I., & Diercks, R. L. (2015). Comparing surgical repair with conservative treatment for degenerative rotator cuff tears: a randomized controlled trial. Journal of Shoulder and Elbow Surgery / American Shoulder and Elbow Surgeons … [et Al.], 24(8), 1274–1281.

21. Lapner, P., Henry, P., Athwal, G. S., Moktar, J., McNeil, D., & MacDonald, P. (2022). Treatment of rotator cuff tears: a systematic review and meta-analysis. Journal of Shoulder and Elbow Surgery / American Shoulder and Elbow Surgeons … [et Al.], 31(3), e120–e129.

22. Leow, J. M., Krahelski, O., Keenan, O. J., Clement, N. D., & McBirnie, J. M. (2022). Functional outcome following arthroscopic repair of massive rotator cuff tears is equal to smaller rotator cuff tears: a retrospective case-control study. Shoulder & Elbow, *14*(1 Suppl), 52–58.

23. Longo, U. G., Carnevale, A., Piergentili, I., Berton, A., Candela, V., Schena, E., & Denaro, V. (2021). Retear rates after rotator cuff surgery: a systematic review and meta-analysis. BMC Musculoskeletal Disorders, 22(1), 749.

24. Lovric, V., Goldberg, M. J., Heuberer, P. R., Oliver, R. A., Stone, D., Laky, B., Page, R. S., & Walsh, W. R. (2018). Suture wear particles cause a significant inflammatory response in a murine synovial airpouch model. Journal of Orthopaedic Surgery and Research, 13(1), 311.

25. Macdermid, J. C., Khadilkar, L., Birmingham, T. B., & Athwal, G. S. (2015). Validity of the QuickDASH in patients with shoulder-related disorders undergoing surgery. The Journal of Orthopaedic and Sports Physical Therapy, 45(1), 25–36.

26. Maher, A., Leigh, W., Young, S., Caughey, W., Hoffman, T., Brick, M., & Caughey, M. (2022). Do age, demographics, and tear characteristics affect outcomes after rotator cuff repair? Results of over 2000 rotator cuff repairs at 5-year follow-up. Orthopaedic Journal of Sports Medicine, 10(8), 23259671221119222.

27. Mandalia, K., Ames, A., Parzick, J. C., Ives, K., Ross, G., & Shah, S. (2023). Social determinants of health influence clinical outcomes of patients undergoing rotator cuff repair: a systematic review. Et Al [Journal of Shoulder and Elbow Surgery], 32(2), 419–434.

28. Mather, R. C., 3rd, Koenig, L., Acevedo, D., Dall, T. M., Gallo, P., Romeo, A., Tongue, J., & Williams, G., Jr. (2013). The societal and economic value of rotator cuff repair. The Journal of Bone and Joint Surgery. American Volume, 95(22), 1993–2000.

29. Mazuquin, B., Moffatt, M., Gill, P., Selfe, J., Rees, J., Drew, S., & Littlewood, C. (2021). Effectiveness of early versus delayed rehabilitation following rotator cuff repair: Systematic review and meta-analyses. PloS One, 16(5), e0252137.

30. Mazzocca, A. D., Arciero, R. A., Shea, K. P., Apostolakos, J. M., Solovyova, O., Gomlinski, G., Wojcik, K. E., Tafuto, V., Stock, H., & Cote, M. P. (2017). The Effect of Early Range of Motion on Quality of Life, Clinical Outcome, and Repair Integrity After Arthroscopic Rotator Cuff Repair. Arthroscopy: The Journal of Arthroscopic & Related Surgery: Official Publication of the Arthroscopy Association of North America and the International Arthroscopy Association, 33(6), 1138–1148.

31. Pastor, T., Zderic, I., Dhillon, M., Gueorguiev, B., Richards, R. G., Pastor, T., & Vögelin, E. (2024). New dynamic suture material for tendon transfer surgeries in the upper extremity – a biomechanical comparative analysis. Archives of Orthopaedic and Trauma Surgery, 144(6), 2905–2914.

32. Rashid, M. S., Cooper, C., Cook, J., Cooper, D., Dakin, S. G., Snelling, S., & Carr, A. J. (2017). Increasing age and tear size reduce rotator cuff repair healing rate at 1 year. Acta Orthopaedica, 88(6), 606–611.

33. Rees, J. L. (2008). The pathogenesis and surgical treatment of tears of the rotator cuff. The Journal of Bone and Joint Surgery. British Volume, 90(7), 827–832.

34. Ren, Y.-M., Zhang, H.-B., Duan, Y.-H., Sun, Y.-B., Yang, T., & Tian, M.-Q. (2019). Comparison of arthroscopic suture-bridge technique and double-row technique for treating rotator cuff tears: A PRISMA meta-analysis. Medicine, 98(20), e15640.

35. Saade, F., van Rooij, F., Saffarini, M., & Godenèche, A. (2023). Management of shoulder stiffness following rotator cuff repair: a systematic review and meta-analysis. *JSES Reviews*, Reports, and Techniques, 3(3), 324–330.

36. Sahoo, S., Stojanovska, M., Imrey, P. B., Jin, Y., Bowles, R. J., Ho, J. C., Iannotti, J. P., Ricchetti, E. T., Spindler, K. P., Derwin, K. A., & Entezari, V. (2021). Changes From Baseline in Patient-Reported Outcomes at 1 Year Versus 2 Years After Rotator Cuff Repair: A Systematic Review and Meta-analysis. The American Journal of Sports Medicine, 3635465211023967.

37. Savage, E., Hurren, C. J., Rajmohan, G. D., Thomas, W., & Page, R. S. (2023). Arthroscopic knots: Suture and knot characterisation of modern polyblend suture materials. Heliyon, 9(9), e19391.

38. Scholes, C., Eng, K., Harrison-Brown, M., Ebrahimi, M., Brown, G., Gill, S., & Page, R. (2023a). Patient Registry of Upper Limb Outcomes (PRULO): a protocol for an orthopaedic clinical quality registry to monitor treatment outcomes. Journal of Surgical Protocols and Research Methodologies, 2023(4), snad014.

39. Scholes, C., Eng, K., Harrison-Brown, M., Ebrahimi, M., Brown, G., Gill, S., & Page, R. (2023b). Patient Registry of Upper Limb Outcomes (PRULO): a protocol for an orthopaedic clinical quality registry to monitor treatment outcomes. Journal of Surgical Protocols and Research Methodologies, 2023(4), snad014.

40. Sheps, D. M., Silveira, A., Beaupre, L., Styles-Tripp, F., Balyk, R., Lalani, A., Glasgow, R., Bergman, J., Bouliane, M., & Shoulder and Upper Extremity Research Group of Edmonton (SURGE). (2019). Early Active Motion Versus Sling Immobilization After Arthroscopic Rotator Cuff Repair: A Randomized Controlled Trial. Arthroscopy: The Journal of Arthroscopic & Related Surgery: Official Publication of the Arthroscopy Association of North America and the International Arthroscopy Association, 35(3), 749–760.e2.

41. Stone, M. A., Henry, T. W., Gutman, M. J., Ho, J. C., & Namdari, S. (2023). Surgical Treatment of Shoulder Infection Following Rotator Cuff Repair. The Archives of Bone and Joint Surgery, 11(2), 111–116.

42. Unger, R. Z., Burnham, J. M., Gammon, L., Malempati, C. S., Jacobs, C. A., & Makhni, E. C. (2019). The responsiveness of patient-reported outcome tools in shoulder surgery is dependent on the underlying pathological condition. The American Journal of Sports Medicine, 47(1), 241–247.

43. Yanik, E. L., Chamberlain, A. M., & Keener, J. D. (2021). Trends in rotator cuff repair rates and comorbidity burden among commercially insured patients younger than the age of 65 years, United States 2007–2016. JSES Reviews, Reports, and Techniques, 1(4), 309–316.

44. Zhao, J., Luo, M., Pan, J., Liang, G., Feng, W., Zeng, L., Yang, W., & Liu, J. (2021). Risk factors affecting rotator cuff retear after arthroscopic repair: a meta-analysis and systematic review. Journal of Shoulder and Elbow Surgery / American Shoulder and Elbow Surgeons … [et Al.], 30(11), 2660–2670.

45. Zwolak, P., Meyer, P., Molnar, L., & Kröber, M. (2022). The functional outcome of arthroscopic rotator cuff repair with double-row knotless vs knot-tying anchors. Archives of Orthopaedic and Traumatic Surgery. Archiv Fur Orthopadische Und Unfall-Chirurgie, 142(1), 25–31.

