## Supplementary 2 for "Low incidence of adverse events or construct failure of a novel high-strength No.2 round suture in rotator cuff repair: An IDEAL Stage 2a assessment retrospective cohort analysis"

### Clinical outcome of no 2 suture (Dynacord)

Corey Scholes, PhD

Chief Science Officer, EBM Analytics

2024-06-13

### 1. Introduction

This analysis links to the manuscript of the dynacord product as one of two companion publications assessing new-to-market hardware. The dataset is derived from the PRULO registry snapshot and live tables. A protocol has been previously prepared for the registry (Scholes et al. 2023).

##### 1.0.1 Preparation

Load up required packages in advance. Citations applied to each library at first use in the text.

1. Load required packages
2. Check if packages are installed, if not, install them

##### 1.0.2 Aim

To describe the clinical and patient-reported outcomes, in patients presenting for surgical review of shoulder pathology and electing to undergo reconstruction or repair of soft-tissue structures with a no. 2 suture (Dynacord, Depuy-Mitek, USA), with or without intracortical anchors at a private, regional orthopaedic clinic between 2020 - 2024.

##### 1.0.3 Hypothesis

No hypotheses have been constructed for this case series as yet.

### 2. Analysis Methods

#### 2.1 Reporting

The study was reported according to the RECORD guidelines (Benchimol et al. 2015) and companion checklist.

The analysis was conducted in RStudio IDE (v2024.04.1+748 “Chocolate Cosmos” Release) using *Rbase* (2023), *quarto* (v1.4) (Allaire and Dervieux 2024) and attached packages to perform the following;

- Data import and preparation
- Sample selection
- Describe missingness
- Data manipulation, modelling and visualisation of;

- Patient characteristics
- Pathology characteristics
- Management and surgical technique
- Treatment and repair survival
- Adverse events and complications
- Patient reported outcomes
  - QDASH
  - WORC Index (Normalised)

#### 3. RECORD [4] - Study Design

Subgroup analysis of a clinical registry embedded into private practice. Observational, cohort design.

#### 4. Data Import and Preparation

Retrieve and format data from live tables and registry snapshot. Using *openxlsx* (Barbone and Garbuszus 2024) to retrieve static snapshot files and *googlesheets4* (v1.1.1) (Bryan 2023) to retrieve live database tables. Text and code output are integrated using the *epoxy* package (v1.0.0) (Aden-Buie 2023).

Read in live tables

Registry snapshot dataframes were combined into one to conduct analysis using *tidyverse* (v2.0.0) (Wickham et al. 2019). Identifiers at the patient level were reconstructed using *stringr* (v1.5.1) (Wickham 2023) functions applied over the dataframe using *purrr* (v1.0.2) (Wickham and Henry 2023). Dates were reformatted to a form appropriate for analysis using *lubridate* (v1.9.3) (Grolemund and Wickham 2011).

#### 5. RECORD [5] - Setting

The PRULO registry is based in a regional private practice for upper limb orthopaedics and has been in operation since July 2020.

The registry has 2681 treatment records with the first patient enrolled 13 October 2020 and the final treatment record created 26 March 2024.The registry snapshot was extracted on 31 March 2024. Patients are followed for up to 2 years after surgery to capture treatment outcomes and patient-reported outcome measures (PROMs).

### 6. RECORD [6] Participants

#### 6.1 RECORD [6.1] Sample selection

Identify cases receiving the suture of interest. Cases were identified by stock keeping units (SKUs) identified from the SKU database maintained as part of implant tracking within the registry. Cases were not restricted by available follow up.

Inclusion criteria;

- Case involves suture of interest
- Case is the index procedure within the registry (first use of suture)
- Patient has not withdrawn consent for inclusion of data in the registry
- Treatment record is eligible for surgery (it has occurred)

Data manipulation (add columns and filter tables based on column values) was performed with *tidyverse* and conversion to display format using *gt* (v0.10.1) (Iannone et al. 2024).

Table 1: Summary of SKUs (Reference) used to identify cases of interest from PRULO registry

| Size (mm) | Description | Category | Reference |
|---|---|---|---|
| 4.5 | Healix Advance BR Dynacord (x2) with Needles | Anchor + Suture | 10886705029440 |
| 4.5 | Healix Advance BR Dynacord (x2) | Anchor + Suture | 10886705029402 |
| 4.5 | Healix Advance BR Dynacord (x3) | Anchor + Suture | 10886705029396 |
| 5.5 | Healix Advance BR Dynacord (x2) | Anchor + Suture | 10886705029464 |
| 5.5 | Healix Advance BR Dynacord (x3) | Anchor + Suture | 10886705029457 |
| 5.5 | Healix Advance BR Dynacord (x2) with Needles | Anchor + Suture | 10886705029471 |
| 6.5 | Healix Advance BR Dynacord (x2) | Anchor + Suture | 10886705029525 |
| 6.5 | Healix Advance BR Dynacord (x3) | Anchor + Suture | 10886705029518 |
| 6.5 | Healix Advance BR Dynacord (x2) with Needles | Anchor + Suture | 10886705029532 |

| Size (mm) | Description | Category | Reference |
|---|---|---|---|
| 4.5 | Healix Advance PEEK Dynacord (x2) with Needles | Anchor + Suture | 10886705029433 |
| 4.5 | Healix Advance PEEK Dynacord (x2) | Anchor + Suture | 10886705029426 |
| 4.5 | Healix Advance PEEK Dynacord (x3) | Anchor + Suture | 10886705029419 |
| 5.5 | Healix Advance PEEK Dynacord (x2) | Anchor + Suture | 10886705029495 |
| 5.5 | Healix Advance PEEK Dynacord (x3) | Anchor + Suture | 10886705029488 |
| 5.5 | Healix Advance PEEK Dynacord (x2) with Needles | Anchor + Suture | 10886705029501 |
| 6.5 | Healix Advance PEEK Dynacord (x2) | Anchor + Suture | 10886705029556 |
| 6.5 | Healix Advance PEEK Dynacord (x3) | Anchor + Suture | 10886705029549 |
| 6.5 | Healix Advance PEEK Dynacord (x2) with Needles | Anchor + Suture | 10886705029563 |
| NA | Gryphon BR Dynacord BL | Anchor + Suture | 10886705029877 |
| NA | Gryphon BR Dynacord STR/BL | Anchor + Suture | 10886705029884 |
| NA | Gryphon P PEEK With Dynacord | Anchor + Suture | 10886705029891 |
| NA | Gryphon P PEEK DS Anchor with Dynacord | Anchor + Suture | 10886705029907 |
| NA | Dynacord #2 suture Pack Blue (with OS-6 needles) | Suture | 222065 |
| NA | Dynacord #2 suture Pack Blue (with MO-7 needles) | Suture | 222066 |
| NA | Dynacord #2 suture Pack Blue (without needles) | Suture | 222067 |
| NA | Dynacord #2 suture Pack Striped (without needles) | Suture | 222068 |

| Size (mm) | Description | Category | Reference |
|---|---|---|---|
| NA | Dynacord #2 suture Pack Striped/Blue (without needles) | Suture | 222069 |
| NA | Dynacord #2 suture Pack Striped/Blue (with MO-7 needles) | Suture | 222071 |
| NA | Dynacord #2 suture Pack Striped/Blue (with OS-6 needles) | Suture | 222073 |

Use SKUs and reference codes to identify treatment records receiving the suture of interest.

Manipulate the registry snapshot to filter any non-index procedures from the master table.

Of the 236 records in the mastersheet, 0 treatment records had withdrawn consent for data inclusion and 2 had declined to participate in PROMs after enrolment.

#### 6.2 RECORD [6.2] Algorithm validation

Registry record selection code was cross-checked by manual record checking within the registry snapshot for a subset (N = 10) of cases.

#### 6.3 RECORD [6.3] Data linkage

No data linkage was utilised for this analysis.

### 7. RECORD [7] Variables

Key variables defined as part of this analysis are summarised in Table 2 below.

Table 2: Summary of key variable definitions in the analysis

| Category | Variable | Comments | Citation |
|---|---|---|---|
| Patient Characteristics | Insurance Status | Recode from account data from practice management system to insurance status | |
| Pathology | CuffRetraction | Defined as per Patte grading | (Lädermann et al. 2016) |
| | CuffCondition | Fatty infiltration as assessed by Goutallier scale | (Fuchs et al. 1999) |
| | TearPattern | Shape the tear makes within the margins of the cuff as viewed in the transverse plane | (Lädermann et al. 2016) |

| Category | Variable | Comments | Citation |
|---|---|---|---|
| | OtherShoulderPathology | Free-text coded as present [Yes] or not [No] | |
| Management - Surgery | RepairAugment | | |
| | CuffTension | | |
| | RepairQuality | | |
| Survival | TreatmentStatus | Labelled as failure after review of clinical notes indicating construct failure (non-operative management) OR reoperation involving removal of index repair hardware | |
| | RetearStatus | Adverse event involving image-confirmed retear or hardware loosening | |
| Adverse Events | Modidifed sink grade | Modification of the Sink grading of complication severity | (Felsch et al. 2021) |
| | Time to event | Time to event for recurring and competing risks | |
| Patient-Reported Outcomes | QuickDASH Total | Total score calculated by summation | (Gummesson, Ward, and Atroshi 2006) |
| | WORC Index Normalised | Total score calculated by summation divided by maximum available score | (Kirkley, Alvarez, and Griffin 2003) |
| | WORC Physical Q3 | How much weakness do you experience in your shoulder? | (Kirkley, Alvarez, and Griffin 2003) |

#### 8. RECORD [8] Data sources

Data was sourced directly from the PRULO clinical registry (Scholes et al. 2023). Patient and treatment information were entered into the database through the registry interface and compiled into a data cube (snapshot) every quarter. Complications and adverse events captured into an online form (QuestionPro, USA) and linked using record identifier codes.

##### 8.1 Adverse Events

Columns in the Complication Table were renamed and joined with the Mastersheet.

Reoperations were identified and subsetted to append to the Complications Table for survival analysis.

Complication entries were written to an external file for co-author review.

The free-text describing the nature of the complication or adverse event was pre-processed using *tidytext* (v0.4.2) (Silge and Robinson 2016) to split into word tokens and remove stop words.

Terms with less than four characters were extracted and reproduced in an external file for manual spelling of abbreviations. Terms with digits (e.g. L5) were removed.

The abbreviated terms with expanded definitions were read back into the workspace for replacement in the complication descriptions.

The terms were replaced and added to the dataframe containing complication data.

A figure displaying term frequency was generated using *ggplot2* (v3.5.1) (Wickham 2016) and formatted for reporting using (v1.47) (Xie 2024).

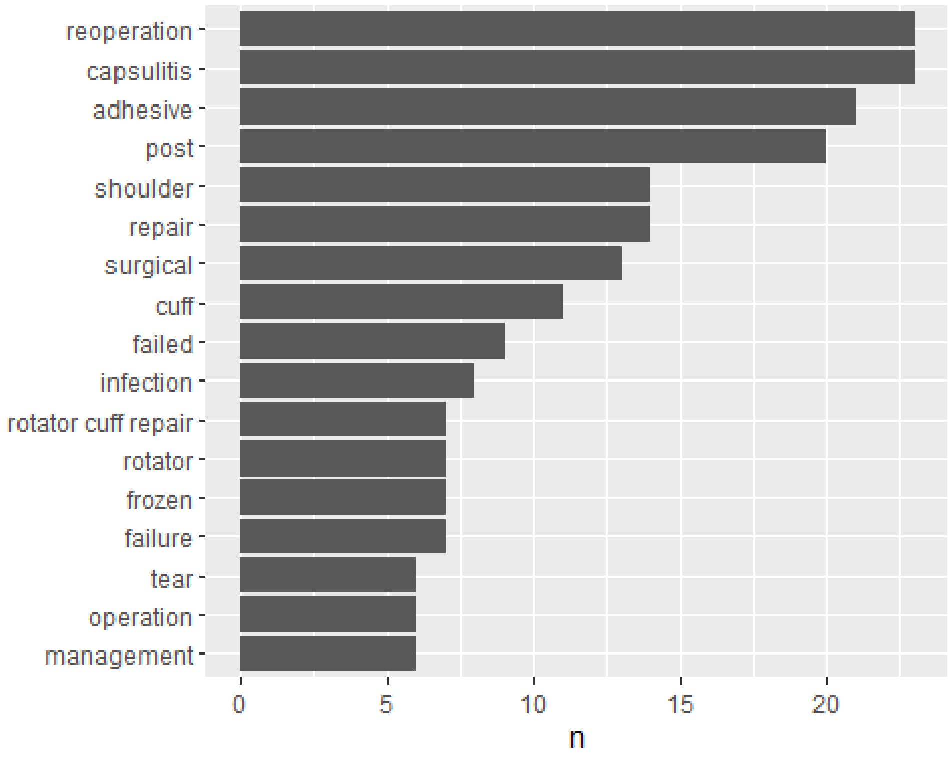

Figure 1: Complication terms by frequency

A wordcloud was generated using *wordcloud* (v2.6) (Fellows 2018) to express the most common terms in the complication description free text field.

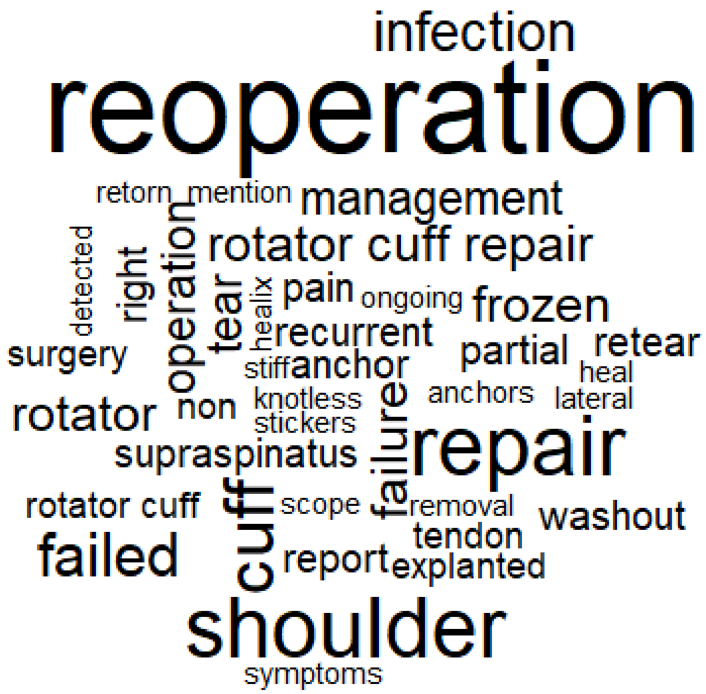

Figure 2: Word cloud of top 50 terms in the complication description data entry

#### 9. RECORD [9] Bias

For a discussion of biases in the context of the clinical registry utilised for this analysis, refer to (Scholes et al. 2023). Specific to this analysis, the following considerations are noted below.

Table 3: Biases in analysis of observational cohort of a clinical registry

| Bias | Definition | Source | Mitigation |
|---|---|---|---|
| Misclassification | Treatment record labelled into incorrect cohort. PROMs package not aligned to | (Benchimol et al. 2015) | Clinical notes reviewed by experienced reviewer and matched to ICD10 code by definition. |
| Confounder | An variable of interest and a target outcome simultaneously influenced by a third variable | (Tennant et al. 2020) | PROMs analysis incorporated adjustment for age and sex |

| Bias | Definition | Source | Mitigation |
|---|---|---|---|
| Missing data | The absence of a data value where a treatment record is eligible to have a data value collected | (Carroll, Morris, and Keogh 2020) | Multiple imputation utilised |
| Prevalent user | Follow-up starts after eligible individuals have started the treatment. The follow-up time is left-truncated | (Nguyen et al. 2021) | Eligibility and enrollment is performed prior to treatment offering for any patient or new presentation. Index procedures identified for analysis are followed prior to surgery occurring. |
| Selection | Treatments are selected based on post-treatment criteria | (Nguyen et al. 2021) | Unable to be mitigated fully - records are identified by presence of hardware code associated with suture of interest |
| Immortal time | Individuals need to meet eligibility criteria that can only be assessed after follow-up has started | (Nguyen et al. 2021) | Patients enrolled at time of diagnosis |
| Pseudoreplication | Analyse data while ignoring dependency between observations. Inadequate model specification. | (Davies and Gray 2015; Lazic 2010) | Cluster for patient in survival (all-cause failure and retear). Utilise mixed effects linear model (lme4::lmer) for PROMs analysis with treatment identifier as random effect |

#### 10. RECORD [10] Sample size

Sample size was derived based on the available records from the Registry at the time of analysis.

#### 11. RECORD [11] Quantitative variables

The anterior-posterior (AP) and mediolateral (ML) dimensions of the cuff tear were reported and multiplied to calculate tear area (mm^2). The tear was also classified according to (Rashid et al. 2017).

- **Small** tears were defined as full-thickness defects in the supraspinatus tendon under 1 cm in the anterior–posterior (AP) dimension.
- **Medium** tears were defined as full-thickness defects in the supraspinatus tendon only, greater than 1 cm and less than 3 cm in the AP dimension.
- **Large** tears involved full-thickness defects of both the supraspinatus and infraspinatus tendons, greater than 3 cm, and less than 5 cm in the AP dimension.
- **Massive** tears involved all 3 tendons (supraspinatus, infraspinatus, and subscapularis) and were greater than 5 cm in the AP dimension.

Partial tears were left labeled as *partial*. Ultimately recoded tear classification based on AP tear length, as the involvement of other tendons for tears of small length was not adequately defined in the original paper.

Retrieved data from the live database table to establish the type of account associated with each treatment record.

Manipulate dataframe - create new coded columns, remove punctuation from free text fields, to provide summary of management and surgery.

Recode free text fields to categorical variables.

The dataset was reshaped to a long format and the indicator columns for each adverse event type were combined into one column within the dataframe (*Category*).

The date of surgery for the index procedure was linked to each complication entry and subsequent quantitative variables such as the durations between;

- date of surgery (index procedure) and date of occurrence
- date of occurrence and date of reoperation
- date of surgery (index procedure) and date of reoperation

Whether the event was intraoperative or presented postoperatively was also assessed using the size of the duration between date of surgery and date of occurrence.

Records with no complication recorded, as well as the final period of right-censor for each record that did not undergo removal of surgery hardware at the end of the chart review period (censored) were generated and added to the complication table to enable reorganisation into a format appropriate for the analysis selected.

Censored treatment records (with no complication recorded at all) were combined with records that were censored after one or more complication events to form the *Censored* component of the adverse events dataset.

The censored data records were integrated into the dataset, with the resultant new frame reorganised into a format appropriate for a multi-state model (see RECORD 12.5) of procedure survival after use of the suture of interest, as described in the *survival* package (v3.6.4) (Therneau 2024).

A *duration* variable was calculated to arrange the dataframe rows within each PatientID in descending order of occurrence to establish the transition patterns from one health state to the next. The start and stop times for certain events (mortality, amputation) were offset by one *week* to remove ties for recurrent events or different event types occurring on the same date for the same patient.The presence of each adverse event type were restricted to the first occurrence of each Category within a patient subsequent to an index procedure per date of occurrence.

The dataframes containing patient-reported outcomes data were manipulated to reduce the columns to those relevant for PROMs analysis and to filter based on eligibility for each time point. The table was subsetted further to analyse the WORC Index (Normalised and Physical Question 3), which was collected without the 3month time point.

### 12. RECORD [12] Statistical methods

A number of analytical techniques were employed to i) clean the data inputs as well as ii) evaluate missingness in the dataset and iii) complete the descriptive analysis of;

- Patient characteristics
- Pathology details
- Patient, implant and adverse event time to event
- Patient-reported outcomes

#### 12.1 RECORD [12.1] Access to population

The registry system represents all cases presenting to the rooms of a surgical group within Geelong, Australia using the implant of interest from the inception of the clinical registry to the analysis date. All reviewed charts from the operating surgeons practice records (electronic medical record) were entered into the database and the present analysis draws data from a regular compilation of the registry records (snapshot) produced quarterly by the registry administration team.

#### 12.2 RECORD [12.2] Data cleaning methods

Diagnosis and complication description free text fields were pre-processed to remove relational terms (stopwords) and expand abbreviations to improve clarity.

Dates of events (preceding and subsequent surgical records; adverse events including mortality) relative to index surgery date were assessed using coded checks to flag anomalies and were resolved by further manual review to resolve inconsistencies or discrepancies with the chart review input data stored in the registry database.

The dataset used as input for the survival analysis of adverse outcomes was assessed survival analysis, with visual assessment of the transitions table to ensure procedure endstates (mortality, implant removal) did not have subsequent states and that the numbers of events and unique identifiers matched the numbers in the dataframe.

```
Call:
survival::survcheck(formula = Surv(DurationStart1, DurationStop1, 
    Category) ~ 1, data = ComplicMaster, id = CombID)

Unique identifiers       Observations        Transitions 
               236                283                 77 

Transitions table:
                to
from              Explant Capsulitis Retear Reoperation ImplantRemoval 
Infection
  (s0)                  4         25     22           0              2   
3
  Explant               0          1      0           2              0   
1
  Capsulitis            0          0      3           1              0   
0
  Retear                0          0      0           0              0   
0
  Reoperation           0          0      0           0              0   
0
  ImplantRemoval        0          0      0           0              0   
0
  Infection             0          0      3           1              0   
0
  Thrombosis            0          0      0           0              0   
1
  Loosening             0          0      0           1              0   
0
  Neurological          0          0      0           0              0   
0
  Other                 0          0      0           0              0   
0
                to
from              Thrombosis Loosening Neurological Other (censored)
  (s0)                     1         0            1     4        174
  Explant                  0         0            0     0          0
  Capsulitis               0         0            0     0         22
  Retear                   0         0            0     0          0
```

```
  Reoperation            0         0            0     0          5
  ImplantRemoval         0         0            0     0          0
  Infection              0         1            0     0          0
  Thrombosis             0         0            0     0          0
  Loosening              0         0            0     0          0
  Neurological           0         0            0     0          1
  Other                  0         0            0     0          4

Number of subjects with 0, 1, ... transitions to each state:
                count
state              0  1 2 3 4
  Explant        232  4 0 0 0
  Capsulitis     210 26 0 0 0
  Retear         208 28 0 0 0
  Reoperation    231  5 0 0 0
  ImplantRemoval 234  2 0 0 0
  Infection      231  5 0 0 0
  Thrombosis     235  1 0 0 0
  Loosening      235  1 0 0 0
  Neurological   235  1 0 0 0
  Other          232  4 0 0 0
  (any)          174 51 8 2 1
```

#### 12.3 RECORD [12.3] Data linkage

Not applicable

#### 12.4 RECORD [12.4] Missingness

##### 12.4.1 Evaluation

Missingness was assessed with visualisation and table functions in the *naniar* package (v1.1.0) (Tierney and Cook 2023) and compiled into figures using *patchwork* (v1.2.0.9000) (Pedersen 2023).

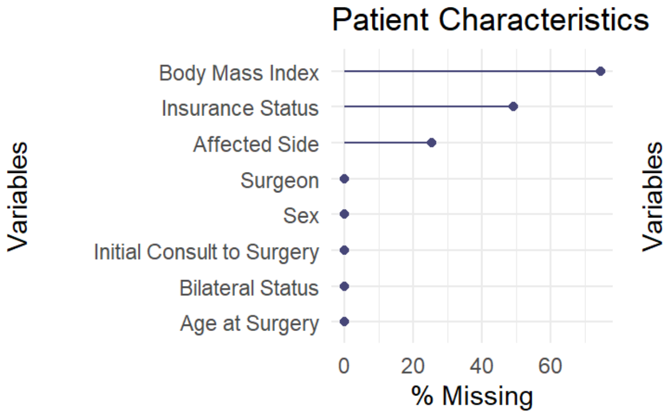
Patient Characteristics
Variables
Body Mass Index
Insurance Status
Affected Side
Surgeon
Sex
Initial Consult to Surgery
Bilateral Status
Age at Surgery
0
20
40
60
% Missing

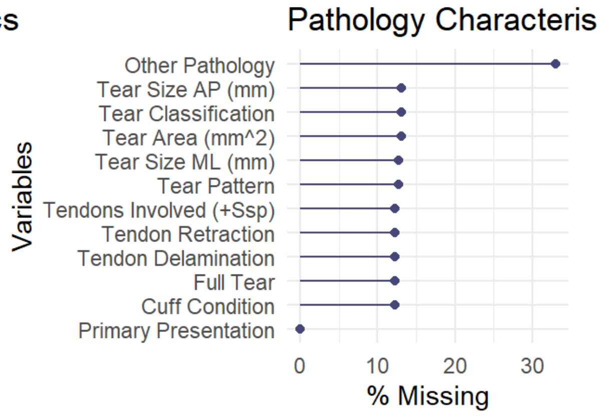
Pathology Characteris
Variables
Other Pathology
Tear Size AP (mm)
Tear Classification
Tear Area (mm^2)
Tear Size ML (mm)
Tear Pattern
Tendons Involved (+Ssp)
Tendon Retraction
Tendon Delamination
Full Tear
Cuff Condition
Primary Presentation
0
10
20
30
% Missing

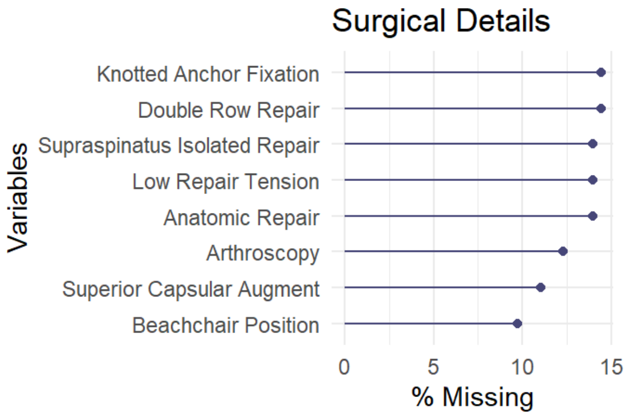
Surgical Details
Variables
Knotted Anchor Fixation
Double Row Repair
Supraspinatus Isolated Repair
Low Repair Tension
Anatomic Repair
Arthroscopy
Superior Capsular Augment
Beachchair Position
0
5
10
15
% Missing

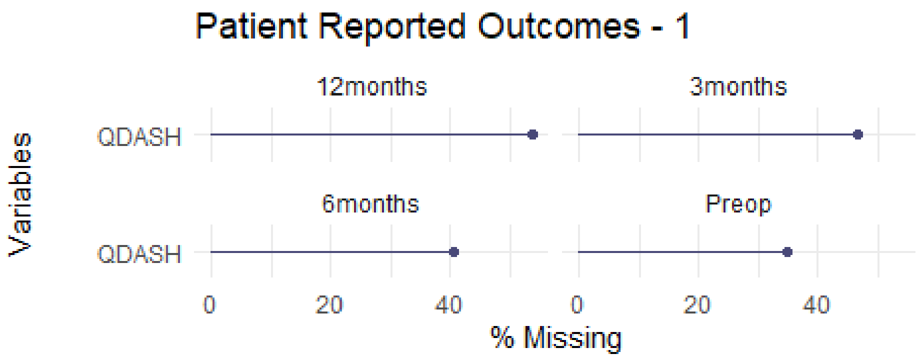
Patient Reported Outcomes - 1
Variables
12months
3months
QDASH
6months
Preop
QDASH
0
20
40
0
20
40
% Missing

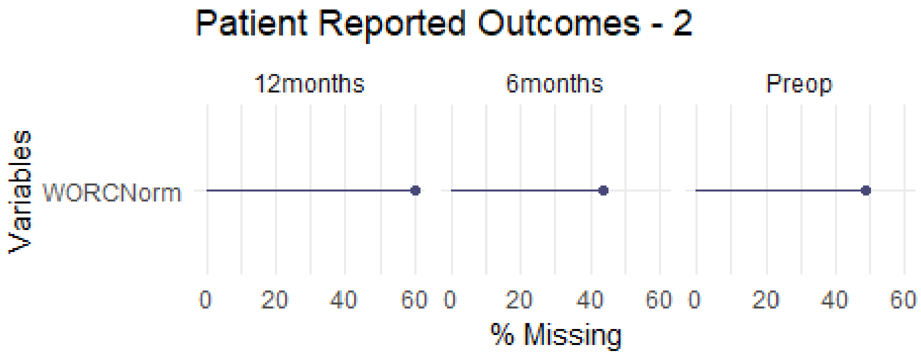
Patient Reported Outcomes - 2
Variables
12months
6months
Preop
WORCNorm
0
20
40
60
0
20
40
60
0
20
40
60
% Missing

Figure 3: Rates of missingness for patient characteristics (A), pathology (B), treatment details (C) and patient-reported outcomes (D and E).

##### 12.4.2 Management

The data tables were sliced to the required columns (PROMs and adjunct columns) in preparation for multiple imputation using chained equations (White, Royston, and Wood 2010) with the *mice* package (v3.16.0) (Buuren and Groothuis-Oudshoorn 2011), with predictive mean matching for continuous variables, logistic regression for binary categorical variables and polytomous logistic regression for ordinal categorical variables.

A density plot (Figure 4) and strip plot (Figure 5) were used to visually inspect the convergence of the imputation iterations against the original dataset.

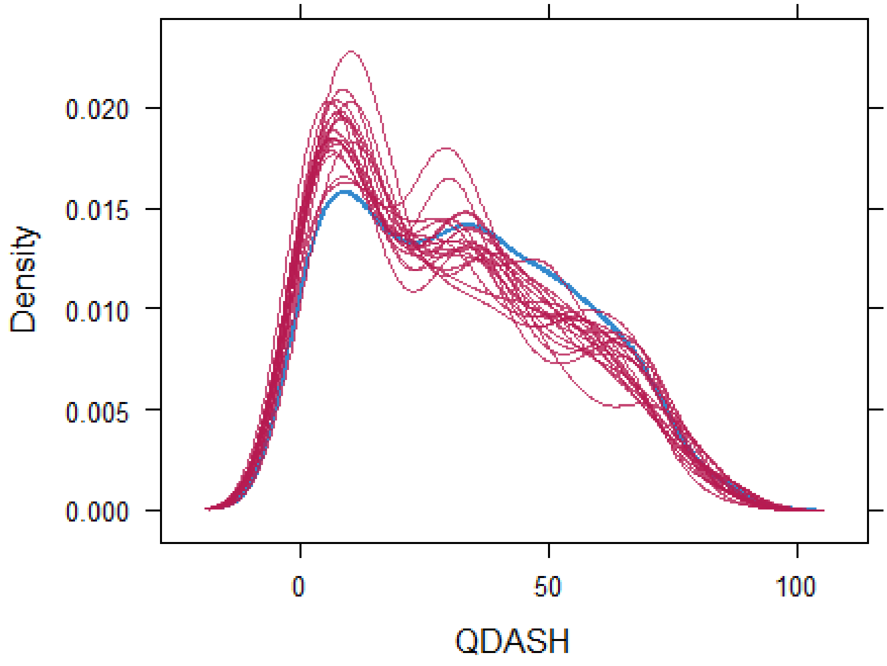

Figure 4: Density plot of QuickDASH imputed over 5 iterations with 20 imputations per iteration against original distribution (blue).

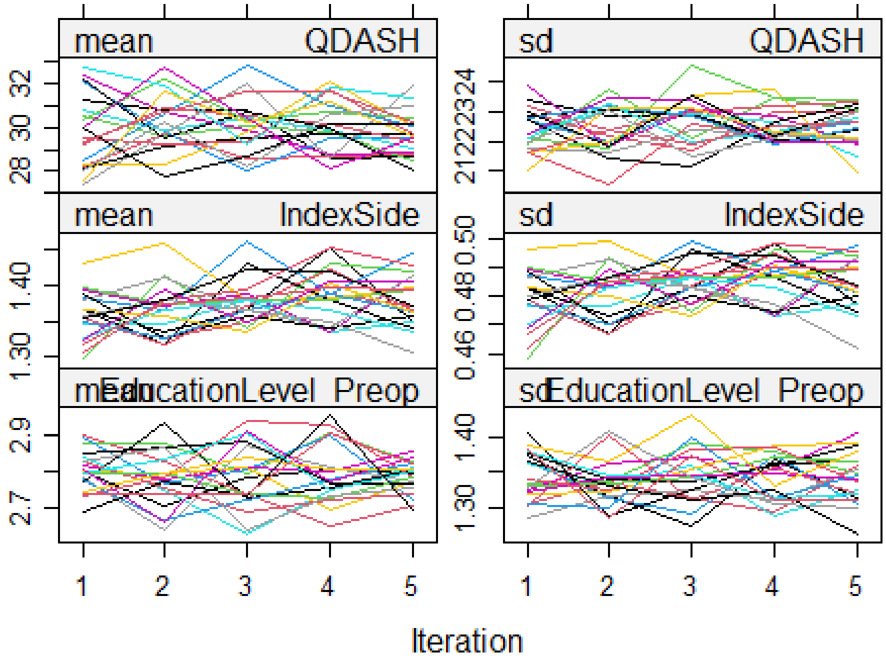

Figure 5: Strip plot of imputed data over 5 iterations with 20 imputations per iteration.

#### 13. RECORD [12.5] Analysis

A flow chart was created with the *consort* package (v1.2.2) (Dayim 2023) to describe the inclusion and exclusion of records into the sample pool for the present analysis to be drawn from. Patient demographics, pathology characteristics and surgical details were summarised using *gtsummary* (v1.7.2) (Sjoberg et al. 2021). Alpha was set for all significance tests at 5%, with confidence intervals of 95% used to bound point estimates for central tendency and model coefficients.

##### 13.1 Adverse Events

The analysis of adverse events and treatment/patient survival after arthroplasty remains a challenging endeavour, made more so by the complexities of ortho-oncology. Attempts have been made to standardize reporting of adverse outcomes after rotator cuff surgery [citations]. However, a key challenge of reporting incidence rates of these outcomes in a given sample is the variability in follow up from one patient to another in the same analytical sample. With variation in followup, the uni-dimensional estimate of incidence (number with condition/total available sample) leads to considerable underestimation of the true rate, since some cases have not yet reached sufficient followup to experience the event of interest. For this reason time-to-event (survival) analysis provides superior incidence estimates - however, there are additional aspects of the present analysis that preclude the use of traditional Kaplan-Meier analysis.

The first is that each patient can experience multiple adverse events after the index procedure (recurring events) which adds a element of dependency to the structure of the adverse event data (Thenmozhi et al. 2019), which is not accounted for in a KM curve. The second is that certain events (e.g. implant removal) preclude the appearance of subsequent adverse events. When these records are subsequently censored (removed from the pool available records) it can bias estimates of other events of interest upward to impossible values (Coemans et al. 2022). In the present dataset, where these elements exist simultaneously, traditional (simplistic) methods can lead to analytical decisions that remove a considerable amount of information from the dataset (e.g. analysis of first occurrence of any type) or biased estimates.

To address these issues within the analysis, the *survival* and *tidycmprsk* (v1.0.0) (Sjoberg and Fei 2023) packages were utilised to deploy a multi-state survival model (see Table 2) to estimate time-varying incidences of competing events such as;

- Implant removal (competing)
- Tendon retear | Hardware breakage
- Infection
- Adhesive capsulitis
- Dislocation - Instability
- Other events

The survival model was expressed in the form;

```
CRModelRCR <- survfit2(Surv(DurationStart1, DurationStop1, Category) ~ 1,
                        data = ComplicMaster,
                        id = CombID
                        )
```

#### 13.2 Patient Reported Outcomes

The QDASH total score and WORC Normalised Index, as well as Question 3 of the Physical sub-scale of the WORC were visualised using the *ggdist* (v3.3.2) (Kay 2023) and *ggplot2* (v3.5.1) (Wickham 2016) packages. Plots were arranged using the *patchwork* (v1.2.0.9000) (Pedersen 2023) package.

Linear regression with random intercepts and timepoint, age and sex as predictors was performed for each PROM scale (QuickDASH, WORC Index Normalised and WORC Q3 Physical) on the multiple imputed datasets using the linear mixed effect model function from *lme4* (v1.1.35.3) , with record identifier included as a random factor. Model-predicted PROM scales were generated using *marginaleffects* (v0.20.1) (Arel-Bundock 2024) and visualised with *ggdist*.

### 14. Analysis Results

#### 14.1 RECORD [13] Participants

The initial export from the registry returned 2681 records of all types.

##### 14.1.1 RECORD [13.1] Treatment selection

The diagram below summarises recruitment and categorisation of patients into the PRULO registry.

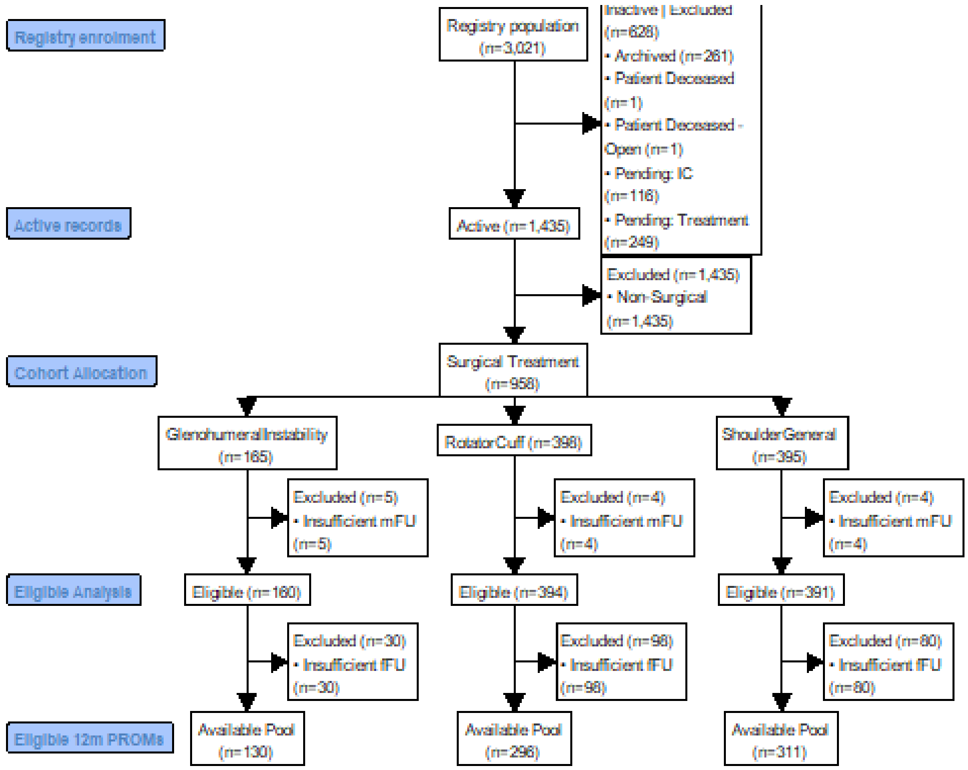

Figure 6: Flow chart of treatment record inclusion and separation into cohorts for analysis. mFU - Minimum follow up (30 days).

#### 14.2 RECORD [14] Patient and record characteristics

Patient characteristics for cases receiving the suture of interest are summarised in Table 4.

Table 4: Summary of patient and treatment record characteristics for the analysed sample.

| Characteristic | N | N = 236 | 95% CI[1] |
|---|---|---|---|
| Age at Surgery, Mean (SD) | 236 | 58 (11) | 57 - 60 |
| Female, % (n) | 236 | 25% (60) | 20 - 32 |

| Characteristic | N | N = 236 | 95% CI[1] |
|---|---|---|---|
| Non-dominant, % (n) | 176 | 36% (63) | 29 - 43 |
| Surgeon, % (n) | 236 | | |
| A | | 53% (126) | |
| B | | 29% (68) | |
| C | | 18% (42) | |
| Bilateral, % (n) | 236 | 8.9% (21) | 5.7 - 13 |
| Registry Cohort, % (n) | 236 | | |
| General | | 0.8% (2) | 0.15 - 3.4 |
| GlenohumeralInstability | | 3.8% (9) | 1.9 - 7.4 |
| RotatorCuff | | 95% (225) | 92 - 98 |
| Exam to surgery delay (weeks), Mean (SD) | 236 | 20 (42) | 14 - 25 |
| Insurance Type, % (n) | 120 | | |
| DVA[2] | | 25% (30) | 18 - 34 |
| Private | | 66% (79) | 57 - 74 |
| TAC[3] | | 2.5% (3) | 0.65 - 7.7 |
| Uninsured | | 6.7% (8) | 3.1 - 13 |
| [1]CI = Confidence Interval | | | |
| [2]DVA = Department of Veterans Affairs | | | |
| [3]TAC = Transport Accident Commission | | | |

The sample was on average, female aged 58 (11) years at the time of surgery with unilateral presentation and privately insured.

##### 14.2.1 RECORD [14.1] Pathology characteristics

Pathology characteristics for cases receiving the suture of interest are summarised in Table 5. The cohort was characterised by majority medium sized full thickness tears in crescent or L shaped of supraspinatus + infraspinatus or isolated supraspinatus with minimal fatty infiltration and tendon retraction.

Table 5: Summary of pathology characteristics for analysed sample

| Characteristic | Available Sample | Summary Statistic | 95% CI[1] |
|---|---|---|---|
| Primary Presentation, % (n) | 236 | 97% (228) | 93 - 98 |
| Full Tear, % (n) | 207 | 92% (190) | 87 - 95 |
| Fatty Infiltration, % (n)[2] | 207 | | |
| 0 | | 54% (112) | 47 - 61 |
| 1 | | 31% (64) | 25 - 38 |
| 2 | | 13% (27) | 8.9 - 19 |
| 3 | | 1.9% (4) | 0.62 - 5.2 |
| Tendon Retraction, % (n)[3] | 207 | | |
| I | | 39% (81) | 33 - 46 |
| II | | 38% (78) | 31 - 45 |
| III | | 15% (31) | 11 - 21 |
| IV | | 7.2% (15) | 4.3 - 12 |
| No retraction | | 1.0% (2) | 0.17 - 3.8 |
| Tendon Delamination, % (n) | 207 | 56% (116) | 49 - 63 |
| Tendons Involved (+Supraspinatus), % (n) | 207 | | |

| Characteristic | Available Sample | Summary Statistic | 95% CI[1] |
|---|---|---|---|
| Infraspinatus | | 14% (29) | 9.7 - 20 |
| Infraspinatus; Subscapularis | | 4.3% (9) | 2.1 - 8.4 |
| Infraspinatus; Teres Minor; Subscapularis | | 0.5% (1) | 0.03 - 3.1 |
| Subscapularis | | 16% (33) | 11 - 22 |
| Subscapularis (isolated) | | 9.7% (20) | 6.1 - 15 |
| Supraspinatus (isolated) | | 56% (115) | 49 - 62 |
| Tear Size AP (mm), Mean (SD) | 205 | 24 (11) | 22 - 25 |
| Tear Size ML (mm), Mean (SD) | 206 | 21 (9) | 20 - 22 |
| Tear Area (mm^2), Mean (SD) | 205 | 560 (489) | 493 - 628 |
| Tear Classification, % (n)[4] | 205 | | |
| Large | | 16% (32) | 11 - 21 |
| Massive | | 1.5% (3) | 0.38 - 4.6 |
| Medium | | 67% (138) | 60 - 74 |
| Partial | | 7.8% (16) | 4.7 - 13 |
| Small | | 7.8% (16) | 4.7 - 13 |
| Tear Pattern, % (n) | 206 | | |
| Crescent | | 45% (93) | 38 - 52 |
| L | | 22% (45) | 17 - 28 |
| Partial articular side | | 3.4% (7) | 1.5 - 7.2 |

| Characteristic | Available Sample | Summary Statistic | 95% CI[1] |
|---|---|---|---|
| Partial bursal side | | 1.9% (4) | 0.62 - 5.2 |
| Reverse L | | 12% (25) | 8.2 - 18 |
| U | | 15% (30) | 10 - 20 |
| V | | 1.0% (2) | 0.17 - 3.8 |
| Other Pathology, % (n) | 158 | 33% (52) | 26 - 41 |
| [1]CI = Confidence Interval | | | |
| [2]Fuchs et al 1999 | | | |
| [3]Modified Patte Grading (Lädermann et al., 2016) | | | |
| [4](Rashid et al., 2017) | | | |

##### 14.2.2 RECORD [14.2] Management summary

Surgical details are summarised in Table 6. Surgery was performed predominantly arthroscopically in beach chair position using anatomic double-row repair of the supraspinatus in isolation using knotted fixation in the majority of cases.

Table 6: Summary of treatment and management details for analysed sample

| Characteristic | Available Sample | Summary Statistic | 95% CI[1] |
|---|---|---|---|
| Arthroscopy, % (n) | 207 | 86% (179) | 81 - 91 |
| Beachchair Position, % (n) | 213 | 86% (183) | 80 - 90 |
| Supraspinatus (isolated) Repair, % (n) | 203 | 59% (119) | 51 - 65 |
| Double Row Repair, % (n) | 202 | 91% (184) | 86 - 94 |
| Knotted Anchor Fixation, % (n) | 202 | 64% (129) | 57 - 70 |
| Superior Capsular Augment, % (n) | 210 | 6.7% (14) | 3.8 - 11 |

| Characteristic | Available Sample | Summary Statistic | 95% CI[1] |
|---|---|---|---|
| Low Repair Tension, % (n) | 203 | 80% (163) | 74 - 85 |
| Anatomic Repair, % (n) | 203 | 90% (183) | 85 - 94 |

[1]CI = Confidence Interval

##### 14.2.3 RECORD [14.3] Follow up

The sample overall had a median follow up of 23 months.

Table 7: Summary of case followup (months) for the sample included for analysis

| Characteristic | **Overall**, N = 236[1] | **Failed**, N = 18[1] | **No further followup**, N = 2[1] | **Ongoing**, N = 216[1] |
|---|---|---|---|---|
| Follow up | 24 (15, 33) | 5 (4, 7) | 10 (7, 13) | 25 (18, 34) |

[1]Median (IQR)

The follow up varied by the type of adverse event observed - as shown below in the Figure.

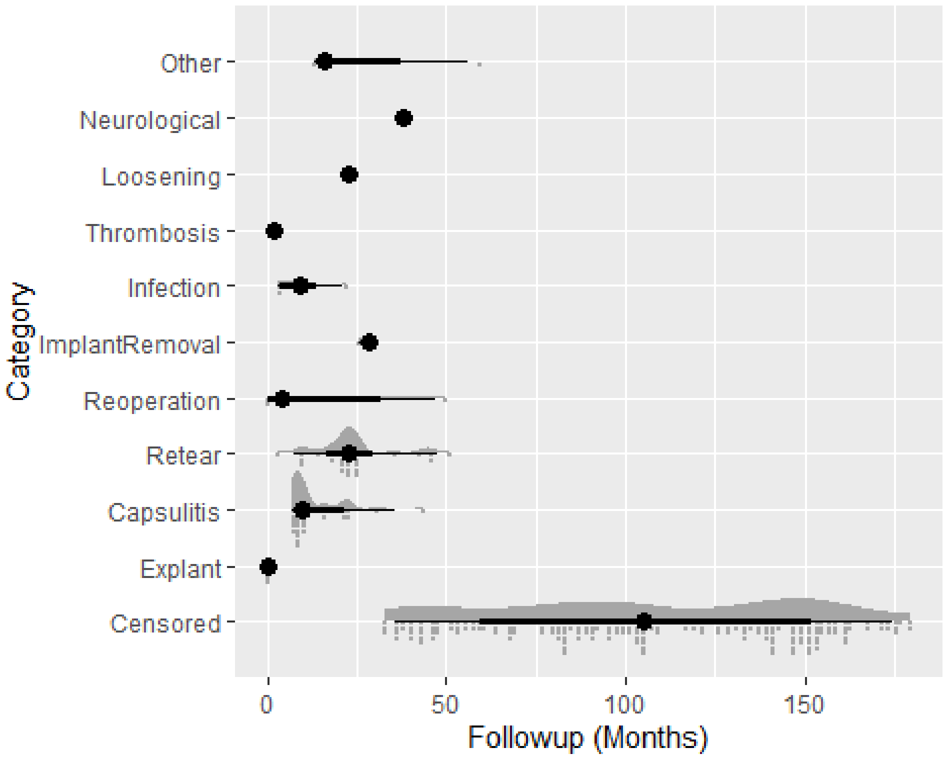

Figure 7: Case follow up separated by adverse event type

#### 14.3 RECORD [15] Outcomes

The outcomes of the present analysis are:

- Adverse events
- Patient-reported outcomes

##### 14.3.1 RECORD [15.1] Adverse events and complications

Table 8: Summary of adverse events after receiving implant of interest (index procedure)

| Characteristic | N = 236 | 95% CI[1] |
|---|---|---|
| Explant, n (%) | 4 (1.7) | 0.54 - 4.6 |
| ImplantRemoval, n (%) | 2 (0.8) | 0.15 - 3.4 |
| Reoperation, n (%) | 5 (2.1) | 0.78 - 5.2 |
| Infection, n (%) | 5 (2.1) | 0.78 - 5.2 |
| Retear, n (%) | 28 (12) | 8.2 - 17 |
| Loosening, n (%) | 1 (0.4) | 0.02 - 2.7 |
| Capsulitis, n (%) | 26 (11) | 7.5 - 16 |
| Neurological, n (%) | 1 (0.4) | 0.02 - 2.7 |
| Thrombosis, n (%) | 1 (0.4) | 0.02 - 2.7 |
| Other, n (%) | 4 (1.7) | 0.54 - 4.6 |
| [1]CI = Confidence Interval | | |

The cohort displayed retear of N = 28 (12)% with (95%CI 8.2 - 17)%, as well as infection (N = 5 (2.1)%, 95%CI 0.78 - 5.2%), implant removal (N = 2 (0.8)%, 95%CI 0.15 - 3.4%) and capsulitis (N = 26 (11)%, 95%CI 7.5 - 16%).

When the adverse events were graded as per Felsch et al, the majority of cases were severity grade I and III. A summary is displayed in Table 9.

Table 9: Cross tabulation of adverse event category with modified Sink grade for complication severity (Felsch et al. 2021)

| Characteristic | **Overall**, N = 69[1] | **I**, N = 29 | **II**, N = 10 | **III**, N = 25 | **IV**, N = 5 |
|---|---|---|---|---|---|
| Category, n (%) | | | | | |
| Capsulitis | 26 (38) | 24 (83) | 1 (10) | 1 (4.0) | 0 (0) |
| ImplantRemoval | 2 (2.9) | 0 (0) | 0 (0) | 2 (8.0) | 0 (0) |
| Infection | 5 (7.2) | 0 (0) | 0 (0) | 5 (20) | 0 (0) |
| Loosening | 1 (1.4) | 0 (0) | 0 (0) | 1 (4.0) | 0 (0) |
| Neurological | 1 (1.4) | 0 (0) | 0 (0) | 0 (0) | 1 (20) |
| Other | 4 (5.8) | 4 (14) | 0 (0) | 0 (0) | 0 (0) |
| Retear | 29 (42) | 1 (3.4) | 8 (80) | 16 (64) | 4 (80) |
| Thrombosis | 1 (1.4) | 0 (0) | 1 (10) | 0 (0) | 0 (0) |

[1]n (%)

Incidence rates were altered when viewed within the context of the multistate survival model. The cumulative incidences, when expressed at set follow up times, showed early peak incidence (<12months of surgery) for infection (Table 10). Cumulative tendon retear also peaked at 20.6% by the 3 year followup.

Table 10: Summary of cumulative incidences of adverse events after rotator cuff repair with suture of interest

| | W4 | | | Wk14 | | | Wk38 | | | Wk52 | | | Wk104 | | | Wk156 | | |
|---|---|---|---|---|---|---|---|---|---|---|---|---|---|---|---|---|---|---|
| | Cum Incid | CI Lower | CI Upper | CumIncid | CI Lower | CI Upper | CumIncid | CI Lower | CI Upper | CumIncid | CI Lower | CI Upper | CumIncid | CI Lower | CI Upper | Cum Incid | CI Lower | CI Upper |
| (s0) | 97.0 | 94.9 | 99.2 | 89.0 | 85.1 | 93.1 | 76.2 | 71.0 | 81.9 | 73.9 | 68.5 | 79.8 | 73.4 | 67.9 | 79.3 | 73.4 | 67.9 | 79.3 |
| Explant | 0.8 | 0.2 | 3.4 | 0.0 | NA | NA | 0.0 | NA | NA | 0.0 | NA | NA | 0.0 | NA | NA | 0.0 | NA | NA |
| Capsulitis | 0.0 | NA | NA | 7.2 | 4.6 | 11.4 | 9.3 | 6.3 | 13.9 | 9.3 | 6.3 | 13.9 | 9.3 | 6.3 | 13.9 | 9.3 | 6.3 | 13.9 |
| Retear | 0.4 | 0.1 | 3.0 | 1.7 | 0.6 | 4.5 | 10.2 | 7.0 | 14.9 | 12.1 | 8.5 | 17.1 | 12.1 | 8.5 | 17.1 | 12.1 | 8.5 | 17.1 |
| Reoperation | 0.8 | 0.2 | 3.4 | 1.3 | 0.4 | 3.9 | 1.7 | 0.6 | 4.5 | 2.1 | 0.9 | 5.1 | 2.1 | 0.9 | 5.1 | 2.1 | 0.9 | 5.1 |
| ImplantRemoval | 0.0 | NA | NA | 0.0 | NA | NA | 0.8 | 0.2 | 3.4 | 0.8 | 0.2 | 3.4 | 0.8 | 0.2 | 3.4 | 0.8 | 0.2 | 3.4 |
| Infection | 0.4 | 0.1 | 3.0 | 0.0 | NA | NA | 0.0 | NA | NA | 0.0 | NA | NA | 0.0 | NA | NA | 0.0 | NA | NA |
| Thrombosis | 0.4 | 0.1 | 3.0 | 0.4 | 0.1 | 3.0 | 0.0 | NA | NA | 0.0 | NA | NA | 0.0 | NA | NA | 0.0 | NA | NA |
| Loosening | 0.0 | NA | NA | 0.0 | NA | NA | 0.0 | NA | NA | 0.0 | NA | NA | 0.0 | NA | NA | 0.0 | NA | NA |
| Neurological | 0.0 | NA | NA | 0.0 | NA | NA | 0.4 | 0.1 | 3.1 | 0.4 | 0.1 | 3.1 | 0.4 | 0.1 | 3.1 | 0.4 | 0.1 | 3.1 |
| Other | 0.0 | NA | NA | 0.4 | 0.1 | 3.0 | 1.3 | 0.4 | 3.9 | 1.3 | 0.4 | 3.9 | 1.8 | 0.7 | 4.7 | 1.8 | 0.7 | 4.7 |

The following figure illustrates the different incidence trajectories for adverse events within each cohort, when taking into account retear and implant removal as competing risks.

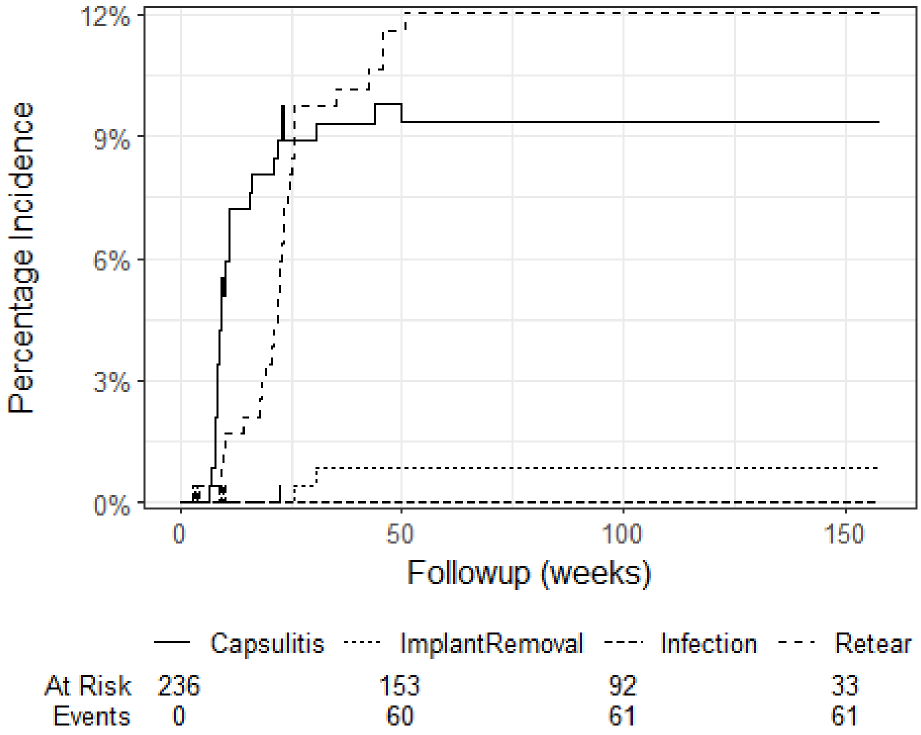

Figure 8: Cumulative incidence of capsulitis in the presence of competing risks (implant removal and tendon retear)

##### 14.3.2 RECORD [15.2] Patient-reported outcome measures

Complete case analysis demonstrated a reduction in QuickDASH total score and WORCPhysicalQ3 and an increase in WORC Normalised from baseline to 12month follow up.

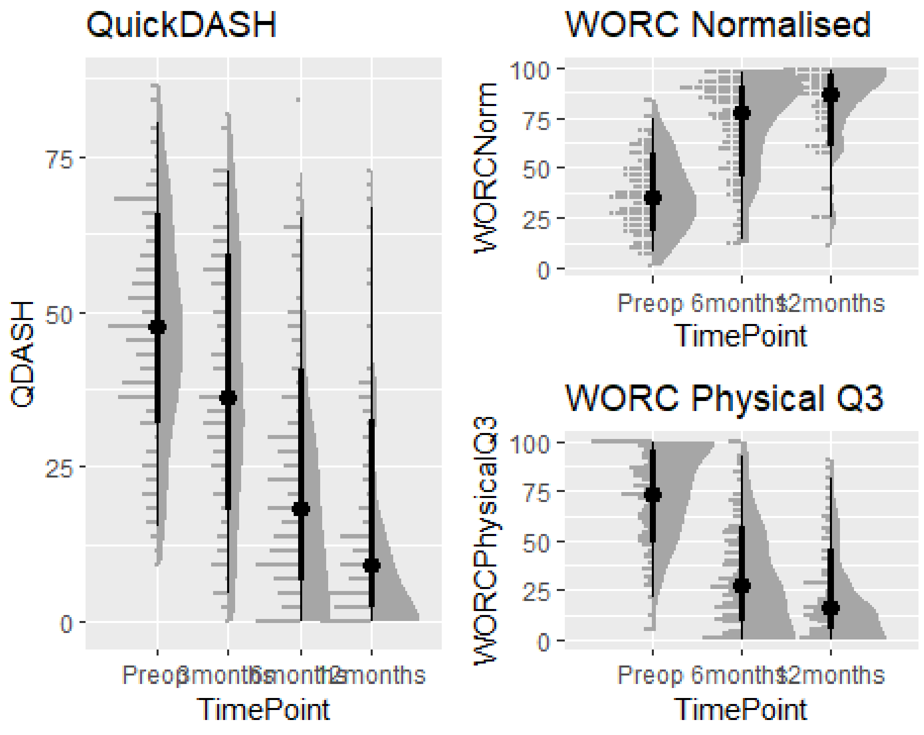

Figure 9: Complete case analysis of QDASH and WORC Index and WORC Q3 Physical by timepoint.

#### 14.4 RECORD [16] Main results

The model displayed a 25 point improvement in QDASH at 6months and 30 points at 12months (Table 11), as well as a 31 point improvement in WORC Index (6 months) and 36 points at 12months.

Table 11: Summary of linear mixed effects model with multiple imputation for QDASH

| Characteristic | Beta | 95% CI[1] | p-value |
|---|---|---|---|
| TimePoint | | | |
| Preop | — | — | |
| 3months | -8.87 | -13.27, -4.47 | <0.001 |
| 6months | -25.04 | -28.85, -21.22 | <0.001 |
| 12months | -31.62 | -35.53, -27.70 | <0.001 |

| Characteristic | Beta | 95% CI[1] | p-value |
|---|---|---|---|
| Age at Surgery | 0.10 | -0.09, 0.29 | 0.306 |
| Male vs Female | 4.79 | -0.09, 9.67 | 0.054 |
| [1]CI = Confidence Interval | | | |

Table 11: Summary of linear mixed effects model with multiple imputation for WORC Index Normalised and WORC Q3 Physical

| | Normalised Index | | | Physical Q3 | | |
|---|---|---|---|---|---|---|
| Characteristic | Beta | 95% CI[1] | p-value | Beta | 95% CI[1] | p-value |
| TimePoint | | | | | | |
| Preop | — | — | | — | — | |
| 6months | 25.31 | 19.81, 30.82 | <0.001 | -29.31 | -35.25, -23.37 | <0.001 |
| 12months | 31.28 | 25.46, 37.10 | <0.001 | -36.31 | -42.83, -29.79 | <0.001 |
| Age at Surgery | 0.26 | -0.01, 0.52 | 0.055 | -0.30 | -0.56, -0.04 | 0.027 |
| Male vs Female | -4.10 | -9.69, 1.49 | 0.149 | 1.60 | -4.93, 8.13 | 0.628 |
| [1]CI = Confidence Interval | | | | | | |

Table 12a: Summary of model-predicted QDASH by TimePoint

| Characteristic | Preop, N = 236[1] | 3months, N = 232[1] | 6months, N = 229[1] | 12months, N = 201[1] |
|---|---|---|---|---|
| QDASH | 48 (36, 61) | 36 (24, 55) | 18 (9, 32) | 11 (2, 25) |
| [1]Median (IQR) | | | | |

Table 12b: Summary of model-predicted WORC by TimePoint

| Characteristic | Preop, **N = 227[1]** | 6months, **N = 220[1]** | 12months, **N = 195[1]** |
|---|---|---|---|
| WORCNorm | 38 (26, 58) | 73 (49, 87) | 82 (61, 92) |

| Characteristic | Preop, **N = 227**[1] | 6months, **N = 220**[1] | 12months, **N = 195**[1] |
|---|---|---|---|
| WORCPhysicalQ3 | 67 (47, 86) | 30 (17, 54) | 20 (9, 50) |
| [1]Median (IQR) | | | |

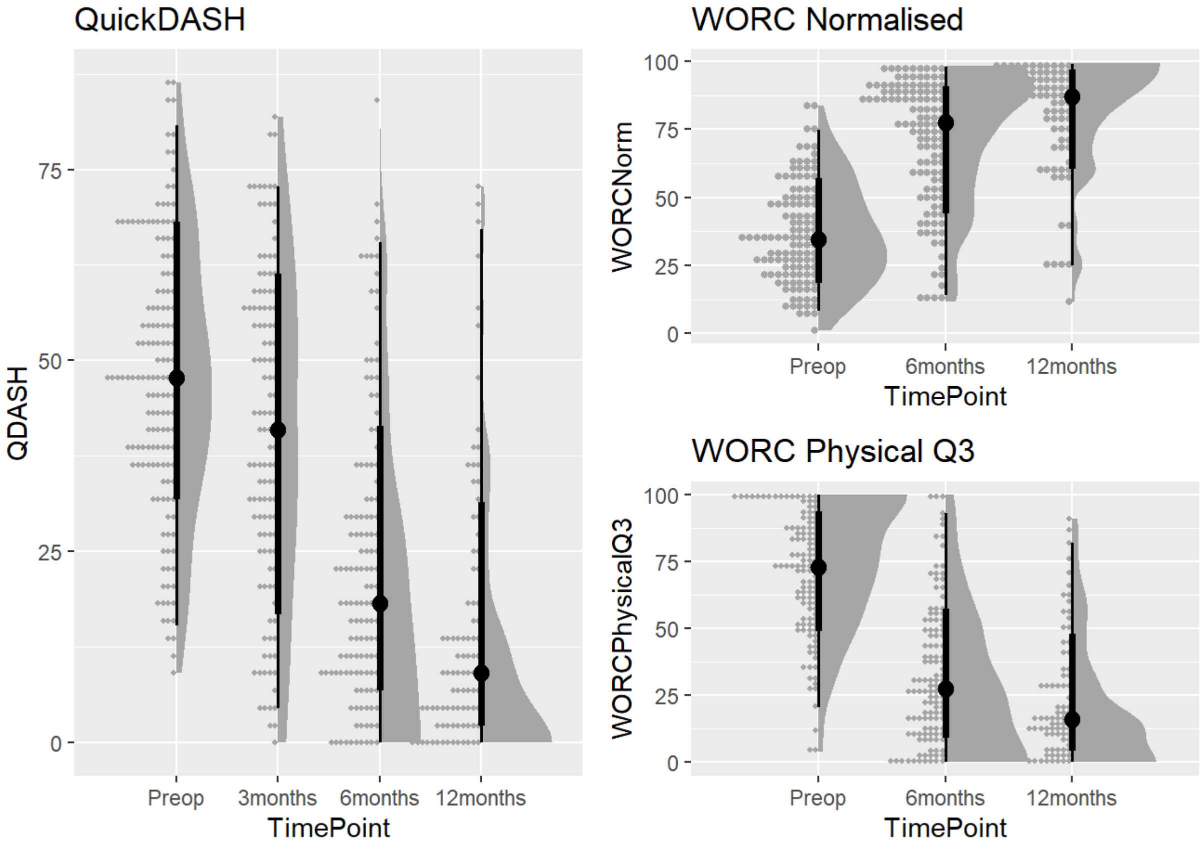

Figure 12: Model predicted trajectories of QDASH (left), WORC Index Normalised (top right) and WORC Q3 Physical (bottom right) across time points

#### 14.5 RECORD [17] Sensitivity analyses

Based on the distribution changes in QDASH and WORC over time, a sensitivity analysis was performed on the model structure using the complete case dataset. A comparison was made between quantile regression using the *quantreg* package (v5.98) (Koenker 2023) and an ordinary least squares linear model from the *rstats* package (v4.4.0) (2022) and a linear mixed effects model with the *lme4* package (v1.1.35.3) (Bates et al. 2015). Results were tabulated using the *modelsummary* package (v2.1.0) (Arel-Bundock 2022) as rq models are not supported in *gtsummary*.

Table 13: Summary of model comparisons between linear fixed effects model, quantile regression and linear mixed effects models

| | RQ | LM | ME |
|---|---|---|---|
| (Intercept) | 38.0 | 41.5 | 40.3 |
| | se = 5.1 | se = 5.0 | se = 6.4 |
| | [28.0, 47.9] | [31.6, 51.3] | [27.6, 53.0] |
| TimePointRecode3months | -9.4 | -8.2 | -9.2 |
| | se = 3.2 | se = 2.2 | se = 1.6 |
| | [-15.6, -3.2] | [-12.5, -3.8] | [-12.4, -5.9] |
| TimePointRecode6months | -27.8 | -24.7 | -25.7 |
| | se = 2.4 | se = 2.2 | se = 1.6 |
| | [-32.6, -23.0] | [-29.0, -20.4] | [-28.8, -22.5] |
| TimePointRecode12months | -37.9 | -32.2 | -30.4 |
| | se = 1.9 | se = 2.4 | se = 1.8 |
| | [-41.7, -34.1] | [-37.0, -27.4] | [-34.0, -26.8] |
| AgeAtTreatment | 0.1 | 0.1 | 0.1 |
| | se = 0.1 | se = 0.1 | se = 0.1 |
| | [0.0, 0.3] | [-0.1, 0.2] | [-0.1, 0.3] |
| Sex2Female | 6.5 | 5.4 | 4.9 |
| | se = 2.0 | se = 1.9 | se = 2.7 |
| | [2.5, 10.4] | [1.6, 9.2] | [-0.4, 10.2] |
| SD (Intercept TreatmentUID) | | | 13.7 |
| | | | se = 1.0 |
| | | | [11.9, 15.8] |
| SD (Observations) | | | 12.8 |
| | | | se = 0.5 |
| | | | [11.8, 13.8] |
| Num.Obs. | 507 | 507 | 507 |
| R2 | 0.299 | 0.324 | |
| R2 Adj. | | 0.317 | |
| R2 Marg. | | | 0.302 |
| R2 Cond. | | | 0.677 |
| AIC | 4422.7 | 4401.8 | 4284.9 |
| BIC | 4448.1 | 4431.4 | 4318.7 |
| ICC | | | 0.5 |
| Log.Lik. | | -2193.907 | |
| F | | 47.918 | |
| RMSE | 18.65 | 18.33 | 10.72 |

The comparison between models revealed an underestimate of the difference in 12month score to preoperative baseline of 7.5 points for the QuickDASH (19.8%) in the quantile regression model, compared to the mixed effects model (random intercept). The mixed effects model also displayed a (42.5%) reduction in root mean square error compared to the quantile regression model.

#### 15. References

Aden-Buie, Garrick. 2023. "Epoxy: String Interpolation for Documents, Reports and Apps." https://CRAN.R-project.org/package=epoxy.

Allaire, JJ, and Christophe Dervieux. 2024. "Quarto: R Interface to 'Quarto' Markdown Publishing System." https://CRAN.R-project.org/package=quarto.

Arel-Bundock, Vincent. 2022. "Modelsummary: Data and Model Summaries in r" 103. https://doi.org/10.18637/jss.v103.i01.

———. 2024. "Marginaleffects: Predictions, Comparisons, Slopes, Marginal Means, and Hypothesis Tests." https://CRAN.R-project.org/package=marginaleffects.

Barbone, Jordan Mark, and Jan Marvin Garbuszus. 2024. "Openxlsx2: Read, Write and Edit 'Xlsx' Files." https://janmarvin.github.io/openxlsx2/.

Bates, Douglas, Martin Machler, Ben Bolker, and Steve Walker. 2015. "Fitting Linear Mixed-Effects Models Using Lme4" 67. https://doi.org/10.18637/jss.v067.i01.

Benchimol, Eric I., Liam Smeeth, Astrid Guttmann, Katie Harron, David Moher, Irene Petersen, Henrik T. Sørensen, Erik von Elm, and Sinéad M. Langan. 2015. "The REporting of Studies Conducted Using Observational Routinely-Collected Health Data (RECORD) Statement." *PLOS Medicine* 12 (10): e1001885. https://doi.org/10.1371/journal.pmed.1001885.

Bryan, Jennifer. 2023. "Googlesheets4: Access Google Sheets Using the Sheets API V4." https://CRAN.R-project.org/package=googlesheets4.

Buuren, Stef van, and Karin Groothuis-Oudshoorn. 2011. "Mice: Multivariate Imputation by Chained Equations in r" 45: 1–67. https://doi.org/10.18637/jss.v045.i03.

Carroll, Orlagh U., Tim P. Morris, and Ruth H. Keogh. 2020. "How Are Missing Data in Covariates Handled in Observational Time-to-Event Studies in Oncology? A Systematic Review." *BMC Medical Research Methodology* 20 (1). https://doi.org/10.1186/s12874-020-01018-7.

Coemans, Maarten, Geert Verbeke, Bernd Döhler, Caner Süsal, and Maarten Naesens. 2022. "Bias by Censoring for Competing Events in Survival Analysis." *BMJ*, September, e071349. https://doi.org/10.1136/bmj-2022-071349.

Davies, G. Matt, and Alan Gray. 2015. "Don't Let Spurious Accusations of Pseudoreplication Limit Our Ability to Learn from Natural Experiments (and Other Messy Kinds of Ecological Monitoring)." *Ecology and Evolution* 5 (22): 5295–5304. https://doi.org/10.1002/ece3.1782.

Dayim, Alim. 2023. "Consort: Create Consort Diagram." https://CRAN.R-project.org/package=consort.

Fellows, Ian. 2018. "Wordcloud: Word Clouds." https://CRAN.R-project.org/package=wordcloud.

Felsch, Quinten, Victoria Mai, Holger Durchholz, Matthias Flury, Maximilian Lenz, Carl Capellen, and Laurent Audigé. 2021. "Complications Within 6 Months After Arthroscopic Rotator Cuff Repair: Registry-Based Evaluation According to a Core Event Set and Severity Grading." *Arthroscopy: The Journal of Arthroscopic & Related Surgery* 37 (1): 50–58. https://doi.org/10.1016/j.arthro.2020.08.010.

Fuchs, Bruno, Dominik Weishaupt, Marco Zanetti, Juerg Hodler, and Christian Gerber. 1999. "Fatty Degeneration of the Muscles of the Rotator Cuff: Assessment by Computed Tomography Versus Magnetic Resonance Imaging." *Journal of Shoulder and Elbow Surgery* 8 (6): 599–605. https://doi.org/10.1016/s1058-2746(99)90097-6.

Grolemund, Garrett, and Hadley Wickham. 2011. "Dates and Times Made Easy with Lubridate" 40. https://www.jstatsoft.org/v40/i03/.

Gummesson, Christina, Michael M Ward, and Isam Atroshi. 2006. "The Shortened Disabilities of the Arm, Shoulder and Hand Questionnaire (Quick DASH): Validity and Reliability Based on Responses Within the Full-Length DASH." *BMC Musculoskeletal Disorders* 7 (1). https://doi.org/10.1186/1471-2474-7-44.

Iannone, Richard, Joe Cheng, Barret Schloerke, Ellis Hughes, Alexandra Lauer, and JooYoung Seo. 2024. "Gt: Easily Create Presentation-Ready Display Tables." https://CRAN.R-project.org/package=gt.

Kay, Matthew. 2023. "Ggdist: Visualizations of Distributions and Uncertainty." https://doi.org/10.5281/zenodo.3879620.

Kirkley, Alexandra, Christine Alvarez, and Sharon Griffin. 2003. "The Development and Evaluation of a Disease-Specific Quality-of-Life Questionnaire for Disorders of the Rotator Cuff: The Western Ontario Rotator Cuff Index." *Clinical Journal of Sport Medicine* 13 (2): 84–92. https://doi.org/10.1097/00042752-200303000-00004.

Koenker, Roger. 2023. "Quantreg: Quantile Regression." https://CRAN.R-project.org/package=quantreg.

Lädermann, Alexandre, Stephen S. Burkhart, Pierre Hoffmeyer, Lionel Neyton, Philippe Collin, Evan Yates, and Patrick J. Denard. 2016. "Classification of Full-Thickness Rotator Cuff Lesions: A Review." *EFORT Open Reviews* 1 (12): 420–30. https://doi.org/10.1302/2058-5241.1.160005.

Lazic, Stanley E. 2010. “The Problem of Pseudoreplication in Neuroscientific Studies: Is It Affecting Your Analysis?” *BMC Neuroscience* 11 (1). https://doi.org/10.1186/1471-2202-11-5.

Nguyen, Van Thu, Mishelle Engleton, Mauricia Davison, Philippe Ravaud, Raphael Porcher, and Isabelle Boutron. 2021. “Risk of Bias in Observational Studies Using Routinely Collected Data of Comparative Effectiveness Research: A Meta-Research Study.” *BMC Medicine* 19 (1). https://doi.org/10.1186/s12916-021-02151-w.

Pedersen, Thomas Lin. 2023. “Patchwork: The Composer of Plots.” https://CRAN.R-project.org/package=patchwork.

R Core Team. 2022. “R: A Language and Environment for Statistical Computing.” https://www.R-project.org/.

Rashid, Mustafa S, Cushla Cooper, Jonathan Cook, David Cooper, Stephanie G Dakin, Sarah Snelling, and Andrew J Carr. 2017. “Increasing Age and Tear Size Reduce Rotator Cuff Repair Healing Rate at 1 Year.” *Acta Orthopaedica* 88 (6): 606–11. https://doi.org/10.1080/17453674.2017.1370844.

Scholes, Corey, Kevin Eng, Meredith Harrison-Brown, Milad Ebrahimi, Graeme Brown, Stephen Gill, and Richard Page. 2023. “Patient Registry of Upper Limb Outcomes (PRULO): A Protocol for an Orthopaedic Clinical Quality Registry to Monitor Treatment Outcomes.” *Journal of Surgical Protocols and Research Methodologies* 2023 (4). https://doi.org/10.1093/jsprm/snad014.

Silge, Julia, and David Robinson. 2016. “Tidytext: Text Mining and Analysis Using Tidy Data Principles in r” 1. https://doi.org/10.21105/joss.00037.

Sjoberg, Daniel D., and Teng Fei. 2023. “Tidycmprsk: Competing Risks Estimation.” https://CRAN.R-project.org/package=tidycmprsk.

Sjoberg, Daniel D., Karissa Whiting, Michael Curry, Jessica A. Lavery, and Joseph Larmarange. 2021. “Reproducible Summary Tables with the Gtsummary Package” 13: 570–80. https://doi.org/10.32614/RJ-2021-053.

Tennant, Peter W G, Eleanor J Murray, Kellyn F Arnold, Laurie Berrie, Matthew P Fox, Sarah C Gadd, Wendy J Harrison, et al. 2020. “Use of Directed Acyclic Graphs (DAGs) to Identify Confounders in Applied Health Research: Review and Recommendations.” *International Journal of Epidemiology* 50 (2): 620–32. https://doi.org/10.1093/ije/dyaa213.

Thenmozhi, Mani, Visalakshi Jeyaseelan, Lakshmanan Jeyaseelan, Rita Isaac, and Rupa Vedantam. 2019. “Survival Analysis in Longitudinal Studies for Recurrent Events: Applications and Challenges.” *Clinical Epidemiology and Global Health* 7 (2): 253–60. https://doi.org/10.1016/j.cegh.2019.01.013.

Therneau, Terry M. 2024. “A Package for Survival Analysis in r.” https://CRAN.R-project.org/package=survival.

Tierney, Nicholas, and Dianne Cook. 2023. “Expanding Tidy Data Principles to Facilitate Missing Data Exploration, Visualization and Assessment of Imputations” 105. https://doi.org/10.18637/jss.v105.i07.

White, Ian R., Patrick Royston, and Angela M. Wood. 2010. “Multiple Imputation Using Chained Equations: Issues and Guidance for Practice.” *Statistics in Medicine* 30 (4): 377–99. https://doi.org/10.1002/sim.4067.

Wickham, Hadley. 2016. “Ggplot2: Elegant Graphics for Data Analysis.” https://ggplot2.tidyverse.org.

Wickham, Hadley. 2023. “Stringr: Simple, Consistent Wrappers for Common String Operations.” https://CRAN.R-project.org/package=stringr.

Wickham, Hadley, Mara Averick, Jennifer Bryan, Winston Chang, Lucy D’Agostino McGowan, Romain François, Garrett Grolemund, et al. 2019. “Welcome to the Tidyverse” 4: 1686. https://doi.org/10.21105/joss.01686.

Wickham, Hadley, and Lionel Henry. 2023. “Purrr: Functional Programming Tools.” https://CRAN.R-project.org/package=purrr.

Xie, Yihui. 2024. “Knitr: A General-Purpose Package for Dynamic Report Generation in r.” https://yihui.org/knitr/.
